# TNFα SIGNALLING IN THE CUTANEOUS IMMUNE NETWORK INSTRUCTS LOCAL Th17 ALLERGEN-SPECIFIC INFLAMMATORY RESPONSES IN ATOPIC DERMATITIS

**DOI:** 10.1101/2021.10.07.21264714

**Authors:** Sofia Sirvent, Andres Vallejo, Emma Corden, Ying Teo, James Davies, Kalum Clayton, Eleanor Seaby, Chester Lai, Sarah Ennis, Rfeef Alyami, Lareb Dean, Matthew Loxham, Sarah Horswill, Eugene Healy, Graham Roberts, Nigel J. Hall, Clare L. Bennett, Peter Friedmann, Harinder Singh, Michael Ardern-Jones, Marta E Polak

**Affiliations:** Systems Immunology Group, Clinical and Experimental Sciences, Faculty of Medicine, University of Southampton; Southampton, UK; University Hospital Southampton NHS Foundation Trust, Southampton, UK; Dermatopharmacology, Clinical and Experimental Sciences, Faculty of Medicine, University of Southampton; Southampton, UK; Department of Haematology, University College London (UCL) Cancer Institute; London WC1E 6DD, UK; Human Development and Health, Faculty of Medicine, University of Southampton; Southampton, UK; BRC, Clinical and Experimental Sciences, Faculty of Medicine, University of Southampton; Southampton, UK; Center for Systems Immunology, Departments of Immunology and Computational and Systems Biology, The University of Pittsburgh; Pittsburgh, USA; Institute for Life Sciences, University of Southampton; Southampton, UK

## Abstract

Accurate regulation of cutaneous immunity is fundamental for human health and quality of life but is severely compromised in inflammatory skin disease. To investigate the molecular crosstalk underpinning tolerance vs inflammation in human skin, we set up a human in vivo allergen challenge study, exposing patients with atopic dermatitis (AD) to house dust mite (HDM). Analyses of transcriptional programmes at the population and single cell levels in parallel with immunophenotyping of resident and infiltrating immune cells indicated that inflammatory responses to HDM were associated with immune activation in Langerhans cells (LCs) and cutaneous T cells. High basal level of TNFα production by cutaneous Th17 T cells predisposed to an inflammatory reaction and resulted in formation of hub structures where LCs and T cells interacted, leading to loss of functional programming in LCs. Additionally, single nucleotide polymorphisms in MT1X gene associated with enhanced expression of metallothioneins and transcriptional programmes encoding antioxidant defences across skin cell types in non-reactive patients, were protective against T cell mediated inflammation. Our results provide a unique insight into the dynamics of immune regulation in the human skin and define regulatory circuits that can be harnessed to improve skin health and treat disease.

## INTRODUCTION

Body surfaces such as the skin, which form the interface with the environment, play a vital role in sensing whether environmental insults are “dangers” and communicate this to the adaptive immune system i^1–5^. Healthy skin is able to maintain immune homeostasis despite of constant environmental insults and an inhabiting biofilm of microbiota. However, in chronic inflammatory skin conditions such as atopic dermatitis (AD), this immunotolerance is breached resulting in uncontrolled immune responses to otherwise innocuous allergens, resulting in flares and exacerbations^6–8^.

AD is one of the most prevalent inflammatory skin conditions, affecting up to 20% of children and 4-7% of adults in European countries^9,10^. 20-30% of AD cases are refractory to treatment, and hence very difficult to manage^11^. Importantly, even though attributing definite causes for eczematous reactions is often impossible, environmental allergens such as pollens, dust mites, pet dander from cats and dogs, moulds and human dandruff, are the commonest triggers inducing allergic immune responses in eczema^8,12^. Such aberrant allergic responses are thought to be a result of a complex crosstalk between an environmental trigger, impaired skin barrier and Th2 adaptive immune activation, resulting in chronic, prolonged inflammation, and uncontrolled flares ^1,13–15^. Interestingly, even though Th2, the key immunophenotype characterising the skin of patients with AD, is overexpressed in the lesional skin, it is also present in the clinically non-inflamed sites ^16,17^, putting in question its definitive role as a sole driver for flares and acute responses.

Advances applying next generation sequencing technologies at the single-cell level resolution progressed our understanding of the complexity of multicellular interactions driving skin disease pathology. Recent research indicates that the regulatory function is dispersed across multiple cell types, cooperating in homeostasis, including immune cells (dendritic cells, T lymphocytes) and parenchyma (keratinocytes, neurons) ^18–20^. While several studies identify that intricate crosstalk between the parenchyma and immune cells is critical for skin homeostasis ^2^, the exact nature of the crosstalk regulating induction of inflammation versus protecting the tolerance and immune homeostasis has not been elucidated.

We sought to delineate the specific interactions within the multicellular cutaneous network underpinning development of cutaneous T-cell mediated immune responses to allergen in skin of patients with AD. A long-standing clinical observation indicates that only a proportion of patients with detectable allergic responses on blood test have positive eczematous responses to local skin challenge with the same allergens. While patients with severe AD show a significantly higher frequency of IgE reactivity to allergens such as cat (Fel d 1) and house dust mite (HDM, Der p 1, 4 and 10) ^21^, it is not understood what determines whether such skin reactivity is present, or why, in some patients with positive blood-derived T cell reactivity to allergens, a skin challenge fails to elicit an eczematous response.

To address this question, we set up a human *in vivo* challenge model exposing patients with AD to a common aeroallergen, house duct mite (HDM), and investigated transcriptional programmes and function of resident and infiltrating immune cells in reactive versus non-reactive patch test sites. This unique approach allowed us not only to delineate a network of interactions in human skin changing dynamically upon exposure to allergen but progressed our understanding of molecular mechanisms safeguarding cutaneous homeostasis. Our analysis indicates that responses to HDM are mediated by Th17 TNF-expressing T cells in reactive patients, driving rapid expansion and overactivation of Langerhans cells (LCs). Interestingly, protection against T cell-mediated inflammation is likely to be conferred by an elaborate network of interactions, including antioxidant responses in the parenchyma. This network contributes to cutaneous non-reactive state, preventing T cell activation and LC exhaustion. It is therefore likely, that therapeutic interventions aimed towards disrupting Th17/TNFα mediated immune cross talk, or directed towards enhancing antioxidant responses, can be harnessed to improve skin health and prevent exacerbations of atopic dermatitis.

## RESULTS

### *In vivo* allergen challenge model to investigate mechanisms of local immune responses in human skin

To investigate the behaviour of systemic and cutaneous human immune systems upon exposure to an allergen, we set up a human *in vivo* allergen challenge study (Figure 1A). We recruited 28 adult patients with moderate to severe atopic dermatitis under the care of a dermatologist in a tertiary referral center. Skin barrier integrity, systemic blood responses and responsiveness to allergen in skin prick test (SPT) were used to assess structural and systemic parameters. The study group comprised 15 males, 13 females, 89% (25/28) of Caucasian ethnicity, with median age = 37 years, (IQR 24.25-53.50), (Supplementary Figure 1A, Supplementary Table 1). Eczema severity scores (EASI) indicated moderate to severe disease (median = 17.7, IQR:10.2 – 30.9, max = 51.4, Supplementary Figure 1A, Supplementary Table 1). Skin barrier was measured as transepidermal water loss (TEWL) of non-eczemtaous sites and was impaired in the AD patients: median = 17.7 g/m^2^h, IQR:13.3 – 30.7, max = 85.0 compared to healthy; median = 8.1 g/m^2^h, IQR = 5.9-10.8, p<0.0001, Supplementary figure 1B). Systemic immune response to HDM was assessed using skin prick test (SPT). 27/28 (96%) patients showed positive SPT reactions to a range of allergens, and the reaction to HDM was one of the strongest (median wheal area 19mm^2^, IQR: 13-26 mm^2^, Supplementary Figure 1C, Supplementary Table 1). All patients had atopic dermatitis (diagnosed by dermatologist as per UK Working Party diagnostic criteria^22^), and the majority suffered with hay fever (24/28, 86%) and asthma (21/28, 75%) (ISAAC questionnaire, Supplementary Figure 1D). Local T cell mediated responses to HDM were measured via *in vivo* allergen exposure patch test and assessed by the clinician. 48 hours post application of a patch test to buttock skin, 11 out of the 28 patients showed clear positive reactions to HDM, and hence, were denoted as “HDM-reactive”. In 12 patients, HDM did not induce a visible response, thus they were labelled as “HDM-non-reactive” (Figure 1B). Four patients reacted to the control patch and were thus labelled as “irritated control reactions”, while 1 patient developed redness in the patch test site which was classified by the clinician as not related to the patch test. This patient was excluded from group analysis.

**Figure 1.**
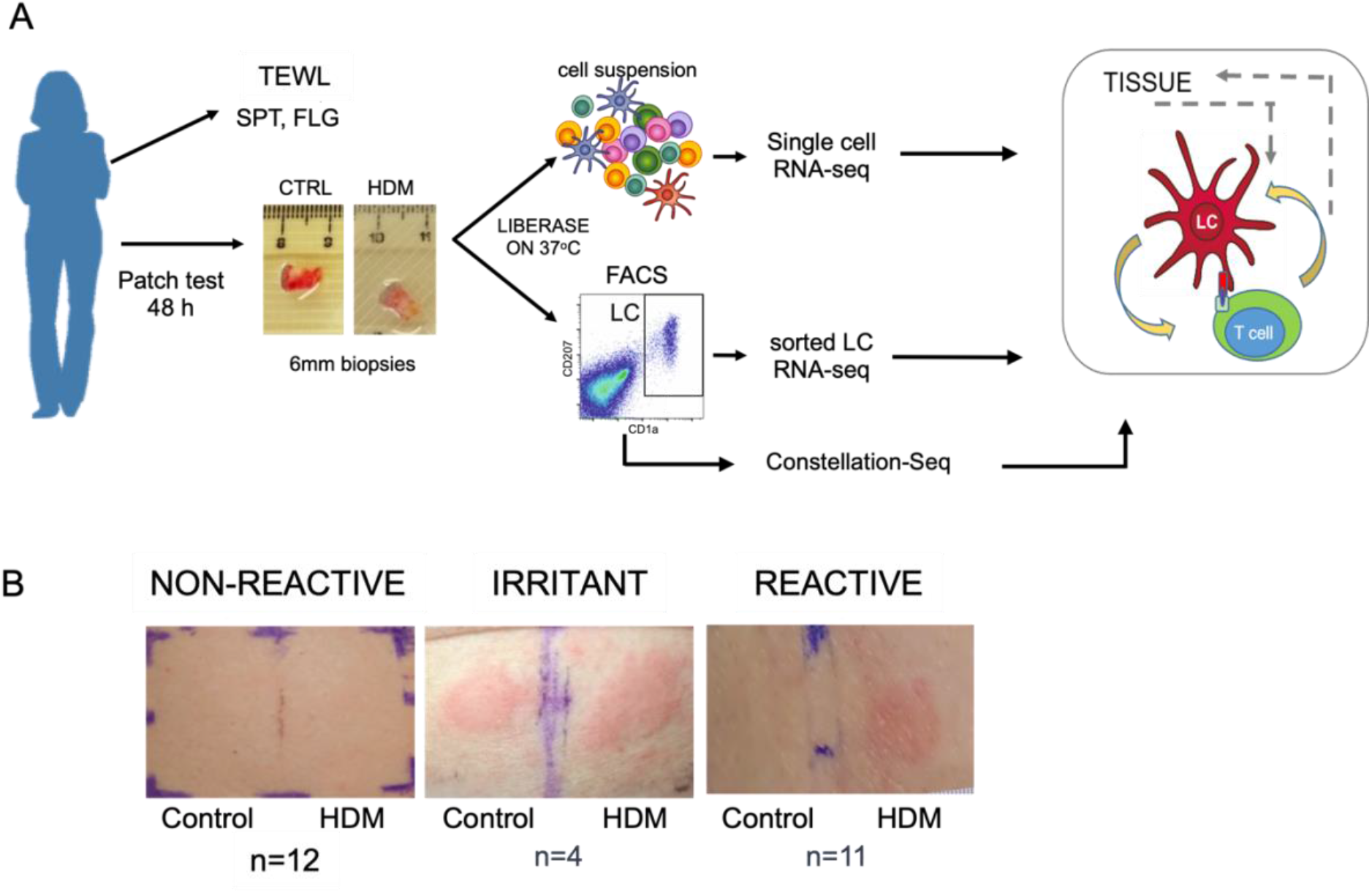
*In vivo* allergen challenge model to investigate mechanisms of local immune responses in human skin. A) Human in vivo allergen challenge set-up. 6mm biopsies taken 48h after application of control (baseline) and HDM (challenge) patch are processed to investigate transcriptional networks and regulatory interactions underpinning T cell mediated responses to allergen (B) Representative images of non-reactive, irritant, and reactive patch test responses to control (left) and HDM (right) allergen, 48 post patch application. Numbers of patients in each group given. TEWL: trans epidermal water loss, SPT: Skin Prick Test, FLG: Filaggrin status

To delineate the cellular and molecular determinants underpinning local cutaneous HDM reactivity versus local tolerance, 6mm punch biopsies were taken from control and HDM patch test sites 48h post *in vivo* challenge with the allergen, representing the baseline and allergen exposed state of skin. To delineate key cell populations, and molecular cross talk pivotal for the T cell-mediated responses developing *in vivo* in human skin we leveraged systems immunology approaches for analysis of next generation transcriptomic and genomic data.

### Reactivity to HDM is mediated by co-expansion of T cells and LCs

One of the possible explanations for the observed lack of responses to HDM in non-reactive patients with known T cell reactivity is a difference in the epidermal permeability barrier, providing better protection against antigen penetration in those patients. However, in our cohort of patients, the measured TEWL level indicated greater epidermal permeability in non-reactive patients (p=0.044, Mann-Whitney test, Figure 2A) Interestingly, high TEWL seemed to predispose to irritated control responses, perhaps highlighting that severely impaired skin barrier facilitates irritant inflammatory reactions. Furthermore, PCR analysis of patient PBMCs showed that the prevalence of mutations in filaggrin (FLG) gene, including 2282del4, R501X, S3247X, R2447X (Fig 2B), was comparable between HDM-reactive and non-reactive patients (p>0.9, chi^2^ test). Subsequently, seven loss of function variants were identified in FLG from whole exome sequencing, validating and extending the PCR results. The additional three variants (Gly1109GlufsTer13, Ser2817AlafsTer75 and Gly323X) were of high quality, had an allele balance >0.19, and did not indicate differences between HDM-reactive and non-reactive patients (p = 0.44, chi^2^ test, Supplementary Figure 2A, Supplementary Table 2). The integrity of transcriptional programming relation to the epidermal barrier from reactive and non-reactive patients was further confirmed using single cell transcriptomic data, indicating programmes encoding tight junctions, desquamation, keratinization, cornification, lipid metabolism and desmosomes were not compromised in the skin of reactive patients (Supplementary Figure 2B, n=6 paired biopsies, Supplementary Table 3).

**Figure 2.**
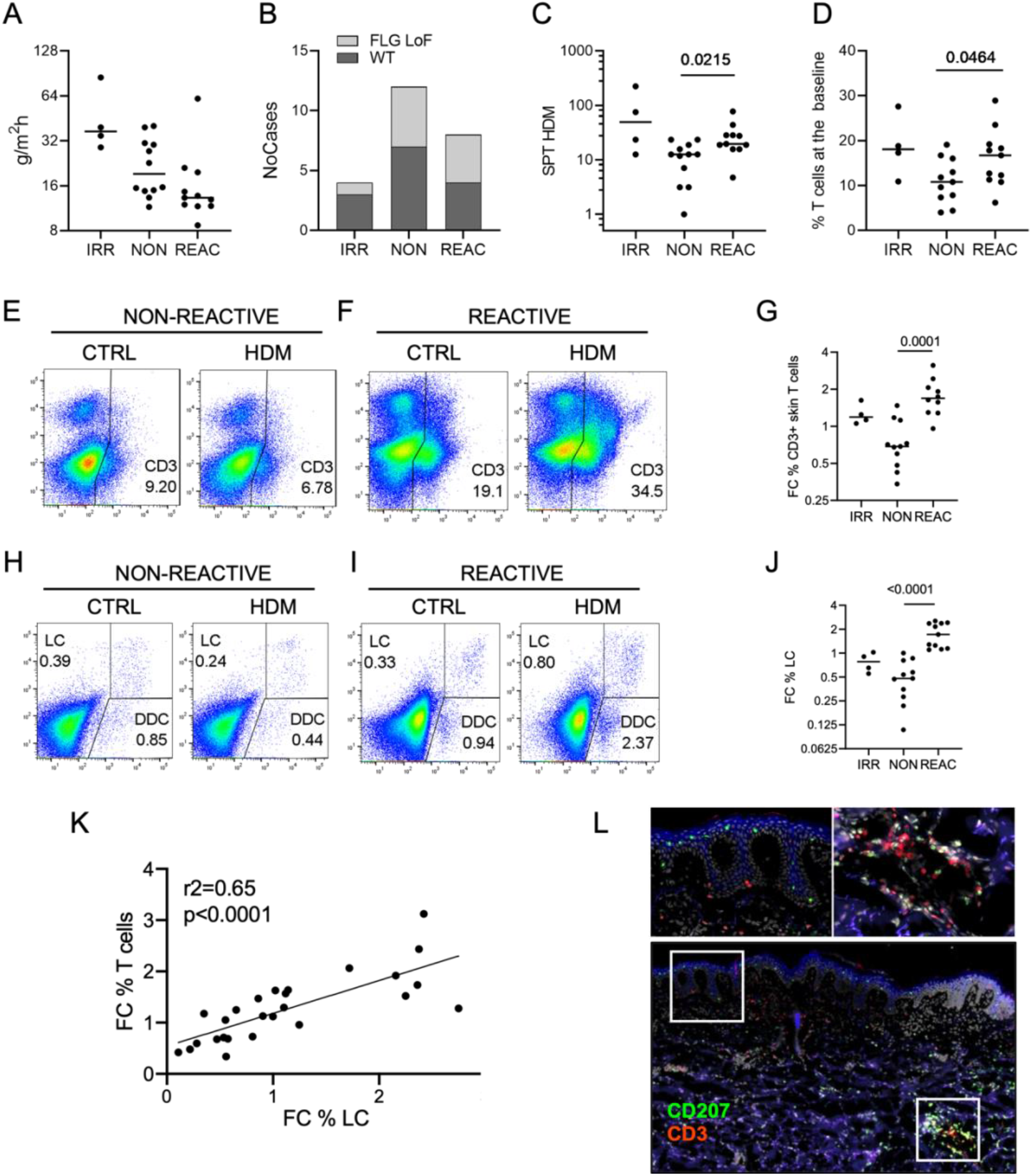
Reactivity to HDM is mediated by co-expansion of T cells and LCs. A) Functional assessment of skin barrier: TEWL measurements across patient groups B) Number of irritant (IRR), non-reactive (NON), and reactive (REAC) cases with loss of function (LoF) variants in FLG compared to wildtype (WT). C) Skin prick test responses to HDM across patient groups, wheal area mm^2^, D) Percentage of CD3+ T cells in control patch test sites identified by flow cytometry. Statistical significance assessed by t-test E-I) Frequency of immune cells in control and HDM patch tests from non-reactive vs reactive patients, measured by flow cytometry. Number in the graph indicates percentage of cells in the positive gate. Representative examples. E-G) CD3+ T lymphocytes, H-J) CD207/CD1a positive LCs. G,I) Fold changes (FC) in percentage of detected immune cells between HDM patch test and control patch test from patients with irritant, non-reactive and reactive reactions to HDM. Statistical significance assessed by t-test J) Correlations between fold changes in percentage of CD3+ T cells and LCs. Pearson correlation coefficient shown. K) Immunofluorescence staining of HDM-reactive patch test site. Inserts show the indicated optical fields at the epidermis (top) and in the dermis (bottom). Hub structures of co-localising CD207 (green) and CD3 (red) in dermis. Epidermal layer stained with multi-cytokeratin (blue). DAPI stain for nuclei (grey).

In contrast to lack of differences in skin barrier function, the wheal areas of HDM-positive SPT were significantly greater in patients with HDM-reactive patch tests (Figure 2C, left panel, p = 0.022, t-test), in agreement with observations by others ^23^. Interestingly, cutaneous CD3+ T cell infiltration at baseline was higher in those patients, possibly indicating a higher immune activation status in skin (Figure 2D, right panel, p = 0.046, t-test). We therefore sought to delineate the immune mechanisms underpinning and coordinating local cutaneous responses to an allergen.

Following HDM application, there was no significant expansion of T cells in non-reactive patch tests or in irritated control reactions (Figure 2E,G), in comparison to the control patch tests. In contrast, in reactive patients, the higher frequency of T lymphocytes at the baseline translated into higher expansion post patch test (p=0.0001, Figure 2F,G). The proportion of HLADR+ cells was visibly higher in responding patients, therefore we sought to determine which population of local antigen presenting cells could be sustaining T cell activation. Quantification of the proportions of LCs (CD207+ CD1a+) versus dermal DC (CD207-CD1a^dim^) in skin biopsies before and after patch testing (Figures 2HIJ, Supplementary Figure 2C) confirmed that a relative expansion of LCs and dermal DCs in reactive, but not tolerant skin compared to baseline.

We observed, a strong correlation between the increase in fold change of LCs and T cells compared to baseline (r^2^=0.59, p<0.0001, Figure 2L) suggesting that immune crosstalk between these cells perpetuates the responses to allergen. In comparison, correlation between dermal DC and T cells was much weaker (Supplementary Figure 2D). In-situ co-localisation of LC (green) and T cells (red) was confirmed in patients reacting to HDM (Figure 2L). Intriguingly, they created hubs akin to tertiary immune structures, inducible skin-associated lymphoid tissue (iSALT) previously described in a mouse model of contact dermatitis ^24,25^. While these structures were less frequent in the control skin of reactive patients, T cells were localised in closer proximity to the epidermis, compared with that of non-reactive patients (Supplementary Figure 2E).

### HDM reactivity in reactive patients is mediated by LC : T cell TNF crosstalk and impairs LC transcriptional programming

The proximity of localisation and co-expansion of LCs and T cells in reactive patch tests suggests that immune crosstalk between these cells is important for T cell mediated cutaneous reaction to HDM. To investigate in depth transcriptional changes in LCs in non-reactive vs reactive patients, CD207+CD1a+ LCs were purified from control and HDM-challenged patch test sites, by fluorescence activated cell sorting (Supplementary Figure 3A), and RNA isolated for whole transcriptome analysis using RNA sequencing. Transcriptome profiles from LCs in control biopsies across patient groups were consistent with steady-state LCs described by us recently ^26^ with programmes encoding cell cycle, including oxidative phosphorylation (BH adj p < 2×10E-8), antigen processing and presentation (BH adj p < 2×10E-8), protein targeting to ER (BH adj p < 2×10E-36) and protein catabolic process (BH adj p = 2×10E-25), (Figure 3A, Supplementary Table 4). Transcript-to-transcript correlation analysis of 28032 filtered and normalised transcripts (BioLayout r=0.85, MCL = 1.7, minimal cluster size = 10 genes) identified 10 clusters of 1115 co-expressed genes, clearly split into two distinct structures (Supplementary Figure 3B). Surprisingly, despite the significant expansion of LC numbers in the reactive HDM-patch test sites, our analyses uncovered impairment of LC transcriptional programming in those samples in comparison to both paired control patch test and to non-reactive patches. The majority of genes (691, Clusters 01,03,04,05,07,08,10) followed a characteristic pattern of expression, with significant downregulation after exposure to HDM in cells isolated from reactive patients (Figure 3B, Supplementary Figure 3C, Supplementary Table 5). This was reflected in downregulation of LCs core transcriptomic programmes, such as protein targeting to ER (BH adj p = 5.67E-82), ubiquitin proteinase ligase binding (BH adj p = 5.83E-10), and antigen processing and presentation (BH adj p =7.38E-9), described by us as key for LC function ^26^. Flow cytometric analyses of LCs across non-reactive and reactive patch-tests indicated that in non-reactive and irritant responses, exposure to HDM increased expression of CD207, the LC hallmark antigen uptake receptor. In contrast, levels of CD207 were markedly decreased in reactive patches, indicating reduction in LCs antigen acquisition and processing function, consistent with observed transcriptional changes (Figure 3D). Unbiased weighted gene expression network analysis (WGCNA,^27^) investigating dependencies between bulk LC transcriptomes and clinical parameters (including TEWL measurements, SPT response, EASI scores), and skin immune parameters (CD3+ T cell infiltration, activation of Th subtypes in blood as measured by flow cytometry) identified 7 modules of co-expressed genes. This unbiased analysis confirmed strong correlation existed between changes of LCs transcriptome modules, disease severity as measured by EASI (module yellow, r=0.48, adj p = 0.001), and level of CD3+ T cell infiltrate in the skin (module turquoise, (|r|=0.31, adj p = 0.04, Supplementary Figure 3D, Supplementary Table 6).

**Figure 3.**
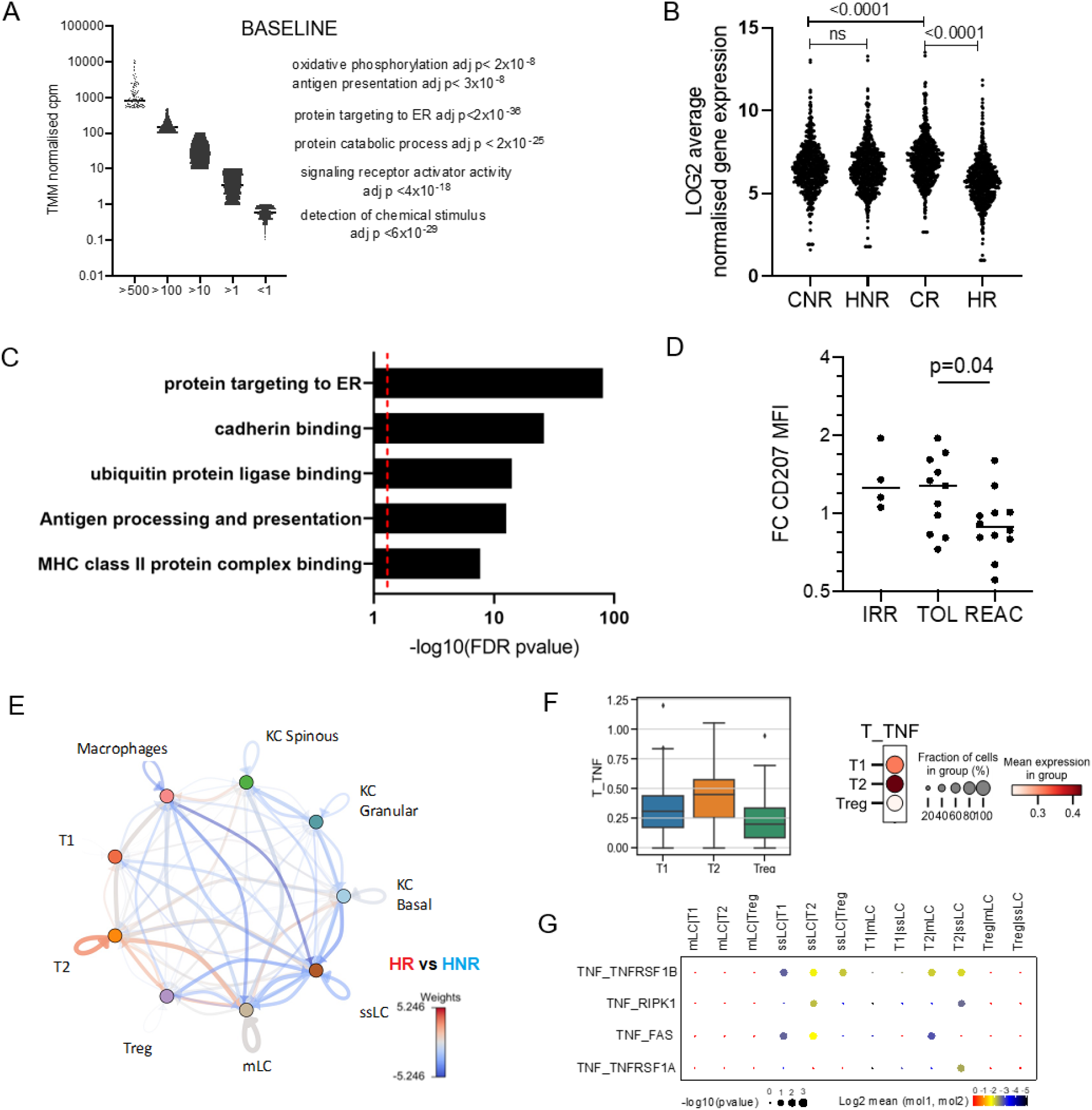
HDM reactivity in reactive patients is mediated by LC : T cell TNF crosstalk and impairs LC transcriptional programming. A) Quantitative transcriptional profile of LC at baseline across all patch tests. Average gene expression binned for the expression level shown. Gene ontology ranked with FDR corrected p-values given for each gene expression level bracket. TMM-normalised counts. B) Average log gene expression levels in the main cluster encoding LC core programmes across non-reactive and reactive patients. Transcript to transcript clustering Biolayout, 691 genes, r=0.85, MCL=1.7. CR: control reactive, HR: HDM reactive, CNR: control non-reactive, HNR: HDM non-reactive C) Gene Ontologies overrepresented in the main cluster encoding LC programmes, TopGene D) HDM-induced change in CD207 expression levels measured by flow cytometry across patient groups n=27, t-test E) Crosstalk analysis of interactions overexpressed between cell populations in non-reactive (blue) and reactive (red) patients after stimulation with HDM. F) TNFα expression across T cell clusters (single cell data, fresh full skin biopsy, n=6 paired biopsies from 3 patients, Drop-seq) G) Dotplot of TNF interactions detected between mature (mLC) and steady state LC (ssLC) and different T cell populations. The size of the dot indicates –log(10) pvalue, the color denotes log(2) average activation strength.

To identify the molecular interactions underpinning LC activation and apparent loss of functional transcriptome, we assessed transcriptional programmes and molecular cross talk in further 6 biopsies from 1 non-reactive and 2 reactive patients using single cell RNA-sequencing. Ligand-receptor analyses of single cell transcriptomes isolated from full skin biopsies confirmed that a signalling edge from activated T lymphocytes delivered activation stimulus via TNF: TNFRSF1B uniquely in HDM-reactive patients (Figure 3E-G). This edge was present at baseline, and became dominant after exposure to HDM (Figure 3E). In contrast, in non-reactive patients, a network of epidermal signalling to LCs was provided by an active crosstalk with multiple cell types, including structural and immune cells.

Interestingly, the same set of genes was differentially expressed at baseline between non-reactive and reactive patients (Figure 3B, p<0.0001), indicating that LCs in reactive patients were more highly activated before exposure to allergen, and pointing to TNFα as the driving stimulus (Supplementary Table 5).

### Activated TNF-expressing Th17 cells are significantly enriched at baseline in reactive patients

Given the observed importance of TNF signalling for HDM responses, we next sought to delineate the specific source population and further delineate the crosstalk programming cutaneous responses to HDM. To reliably track rare populations including specific T cell types, and transcriptional programmes at the level of transcription factors within dissociated skin biopsies we applied Constellation-seq^28^, a highly sensitive transcriptome read-out (Supplementary Table 7). Constellation-seq analysis identified 6 distinct T cell populations, annotated as CD4 naïve T cell, CD4 follicular helper T cells (Tfh), CD8 cytotoxic T cells, primed T cells, regulatory T cells (Tregs), and a small cluster of T cells, using Human Cell Atlas signatures^29^ (Figure 4A,B).

**Figure 4.**
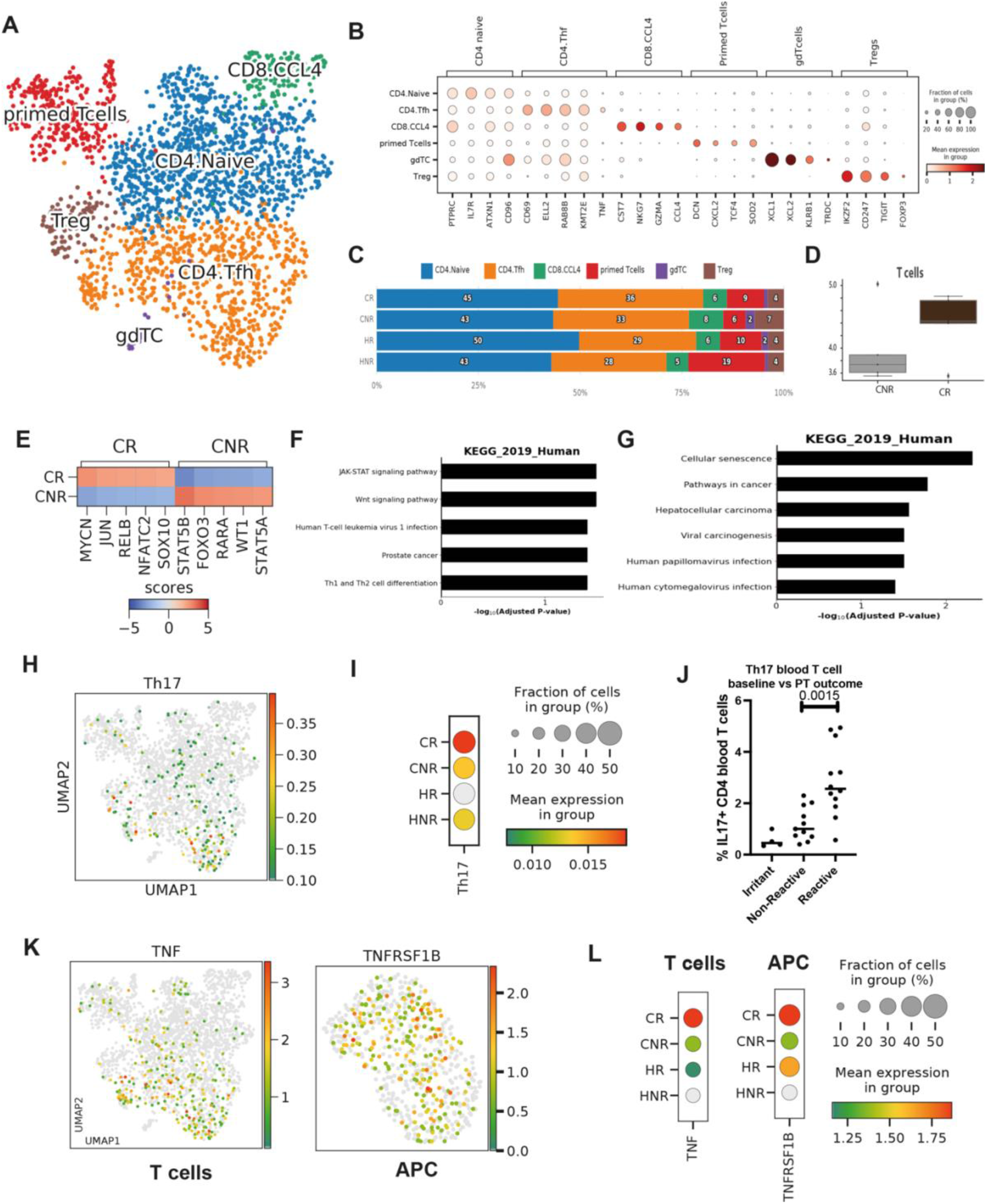
Activated TNF-expressing Th17 cells are significantly enriched at baseline in reactive patients. Constellation-seq analysis enriched for 1161 transcripts in 2374 single T lymphocytes cells from patch test skin biopsies, n=10 patients, 5 per group A) UMAP plot depicting clustering of T lymphocyte populations B) Cell subset defining markers (Wilcoxon rank test). C) T cell subset composition across patient groups D) Number of T cell transcriptomes in control non-reactive (CNR) and reactive (CR) patients E) Top transcription factors expressed in T cells from reactive (CR) and non-reactive (CNR) patients at baseline F,G) Top biological pathways enriched at baseline in DEGs from (F) patient reactive (CR) and (G) non-reactive (CNR) to HDM (KEGG database) H) UMAP plots showing expression of Th17 gene signature I) Th17 gene signature across patient groups, dotplot: size depict % of expressing cells, colour intensity encodes mean expression in group. J) %IL17 producing CD3+CD4+ Tcells from PBMCS in non-reactive (CNR) and reactive (R) patients. T-test. K) UMAP plots showing expression of *TNF* and *TNFRSF1B* across T cell (left) and APCs (right) L) *TNF* and *TNFRSF1B* expression level across patient groups, dotplot: size depict % of expressing cells, colour intensity encodes mean expression in group. CR: control reactive, HR: HDM reactive, CNR: control non-reactive, HNR: HDM non-reactive

Comparative analyses of the prevalence and activation of specific T cell clusters across patient groups showed that T cell changes in response to allergen were quantitative rather than qualitative (Figure 4C), as cell numbers at baseline differed significantly between reactive and non-reactive patients (Figure 4D). Together with changes in T cell numbers observed by flow cytometry (Figure 2G), this strongly indicated that the inflammatory process in HDM patch test site is driven by T cell expansion from populations present at baseline. Interestingly, a trend was observed for HDM-induced expansion of Tregs in patients clinically non-responding to HDM (Supplementary Figure 3A). Contrasting T cell populations in reactive and non-reactive patients indicated that in reactive patients higher activation status of T cells was seen across clusters, manifested by up-regulation of *NFATC2, JUN* and NFkB transcription factors (as exemplified by *RELB*, Figure 3E). In contrast, T cells from non-reactive patients expressed transcription factors regulating tolerogenic properties (*STAT5B*) (Figure 3E). Consistently, genes up-regulated in reactive patients encoded T cell activation via JAK-STAT and Wnt signalling pathways (Figure 3F). In contrast, in patients non-reactive to HDM, T cells showed enrichment of processes of cellular senescence (Figure 3G).

Unbiased clustering analysis revealed statistically significant overrepresentation of TNF expressing T cells within Tfh cluster (Figure 4A,K, TWEAK signaling pathway enriched in CR vs CNR, adj p value 0.019) in unstimulated skin of patients reactive to HDM. Testing for specific T cell transcriptional programmes including Th1, Th2, Th17, Th22, and Treg (as defined by HCA), confirmed enrichment in Th17 cells, overrepresented across Tfh and CD4 naïve clusters, co-localising with TNF-expressing T cells. These cells were highly activated at baseline, delineating a specific immunophenotype of CD3+IL17+TNF+CD69+ T cells overrepresented at baseline in the skin of patients reactive to HDM.

In contrast, Th1, Th2 and Th22 immunophenotypes were shared between cells across patient groups and T cell populations, resembling continuous phenotype clouds of T cells in gut tissue, as described by Kiner an colleagues^30^. Interestingly, even though Th2 determinants were expressed more strongly in patients with reactive HDM-patch tests, Th2 polarisation, albeit weaker, was also evident in non-reactive patients, both at baseline and following *in vivo* challenge with the allergen (Supplementary Figure 4B,C), in agreement with earlier findings reporting Th2 responses in non-lesional eczema skin^16,17^.

To test, whether the Th17 overexpression was translated into functional protein synthesis, we assayed cytokine expression in peripheral blood mononuclear cells from reactive and non-reactive patients. While no differences were discovered in IL13-producing CD4+ T cells between patient groups (Supplementary Figure 4C), IL-17 producing T cells were significantly overrepresented in blood of reactive patients even prior to stimulation with HDM (Figure 4J). The importance of TNF for inter-cellular communication in the skin of reactive patients was next confirmed in crosstalk analyses, demonstrating a TNF: TNFSFRB signaling edge between T cells and APCs, already present at baseline, and strengthened on exposure to HDM in reactive patients.

### Sub-clinical inflammation and environmental stress in differentiated keratinocytes poise immune status of reactive patients

Next, we sought to delineate the cellular and molecular drivers of activated T cell status in the skin. Scanpy analysis ^31^ of Constellation-seq data of 25844 cells filtered for number of expressed genes and viability as assessed by mitochondrial gene content (Supplementary Figure 5A), identified 15 major cell clusters, representing cell populations found in skin biopsies (Leiden algorithm, r=0.5, Figure 5A, Supplementary Table 8). Cell identity was confirmed using HCA marker genes^29^ (Figure 5B). Distinct clusters grouped keratinocytes (undifferentiated and differentiated), immune cells (including APCs and T cells), four populations of fibroblasts (F1-F4), vascular endothelial cells, lymphatic endothelial cells and pericytes and melanocytes (Supplementary Table 8).

**Figure 5.**
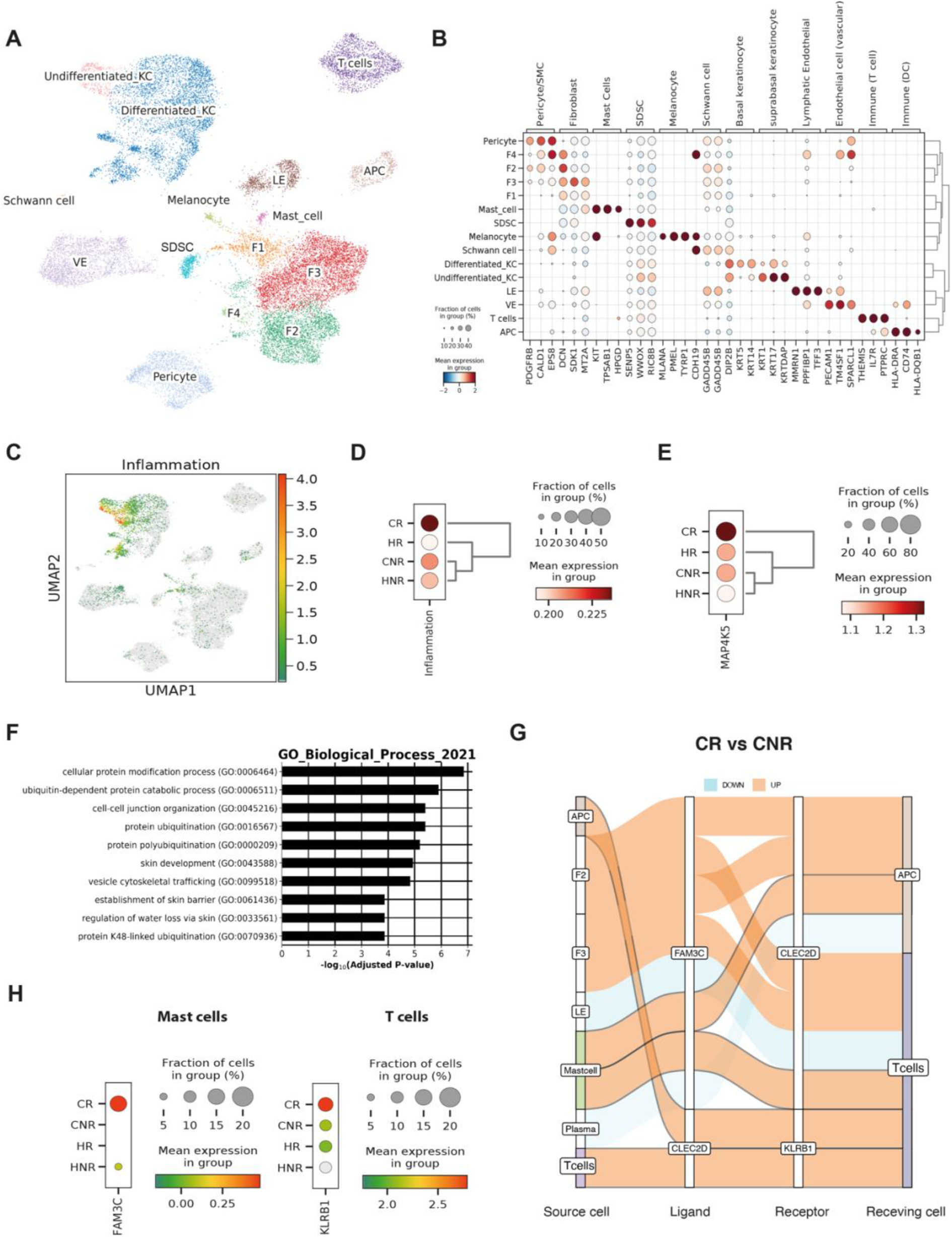
Sub-clinical inflammation drives responses to allergen. Constellation-seq analysis enriched for 1161 transcripts in 25844 single cells from patch test skin biopsies, n=10 patients A) UMAP plot depicting clustering of specific cell populations B) Cell subset defining markers (Wilcoxon rank test). C,D) Expression of inflammation signatures in differentiated keratinocytes. E) Expression of MAP4K5 in differentiated keratinocytes across patient groups. F) Top pathways overrepresented at baseline in differentiated keratinocytes from reactive patients. G) Sankey plot listing all interactions associated with FAM3C and CLEC2D. H) Dotplot demonstrating changes in the expression levels on source and receiving cells among the experimental groups. dotplot: size depict % of expressing cells, colour intensity encodes mean expression in group. CR: control reactive, HR: HDM reactive, CNR: control non-reactive, HNR: HDM non-reactive

Differential gene expression analyses performed for each specific cell population between sample phenotypes indicated that amongst all the cell populations the most DEGs differentiating reactive and non-reactive patients were expressed at baseline by differentiated KCs (825 DEGs, MAST, FDR<0.05, Supplementary table 8). These encoded skin differentiation (FDR p=6×10E-13), hyperkeratosis (FDR p=4×10E-3) and skin inflammation (FDR p=3×10E-3 (ToppGene, Supplementary table 9). Testing for enrichment of pre-defined signatures confirmed that in reactive patients, there was enhanced inflammatory status of differentiated keratinocytes at baseline (Figure 5C,D), which was validated by unbiased single cell whole transcriptome analysis and could be localised to the spinous layer (Supplementary Figure 5 D,E). The activated immune status was epitomised by kinase activity (Supplementray table 9). Interestingly, one of the most highly overexpressed genes, *MAP4K5*, has been implicated in responses to environmental stress (Figure 5E).

Even though very few differentially expressed genes were found in other cell populations at baseline, analysis of molecular crosstalk indicated their importance in signalling to T lymphocytes. A signalling edge between FAM3C on mast cells to CLEC2D was associated with driving T cell maturation and activation to terminal differentiation, demonstrated by KLRB1 expression (Figure 5G,H). Importantly, activation via CLEC2D has been shown to drive TNF expression in multiple cell types ^32^ providing a plausible explanation for TNF-expressing Tfh T lymphocytes observed at baseline in HDM-reactive patients.

While differences at baseline localised specifically to epidermis, in reactive patients exposure to HDM induced activation across many more skin cell types, including fibroblasts and venous endothelial cells, which corresponded to the inflamed, lesion status of the patch test (Supplementary Figure 5, Supplementary Table 10). Importantly, after the exposure to HDM, differentiated keratinocytes initiated HIF1 signalling pathway, providing further evidence that the cutaneous inflammation was associated with hypoxia and oxidative stress (Supplementary Figure 5).

### Enhanced expression of metallothionein genes protects non-reactive patients from HDM-induced T cell-mediated inflammation

Having identified that baseline epidermal sub-clinical inflammation, high activation status of the cutaneous immune compartment, and TNF signalling predisposed to positive inflammatory responses to HDM, we sought to evaluate the parameters protective against inflammation and conducive to local immunotolerance. While analyses of differentially expressed genes across different skin populations identified only sporadic genes up-regulated in non-reactive patients, these differentially up-regulated genes consistently included members of metallothionein family, with *MT2A* being the top overexpressed gene in non-reactive fibroblasts and venous endothelium (FDR p =0.015, supplementary Figure 6A). Drop-seq analyses of freshly dissociated biopsies (n=6) confirmed the high expression of metallothionein gene family across cell populations in the non-reactive vs reactive patients (Figure 6A,B). To ensure that the detection limits and drop-outs were not masking metallothionein expression we further corroborated the results using high-sensitivity Constellation-seq (Figure 6C). While expression of metallothionein transcripts was reduced on exposure to HDM in all patients, cells from biopsies non-reactive to HDM retained some expression of metallothioneins (Figure 6C).

**Figure 6.**
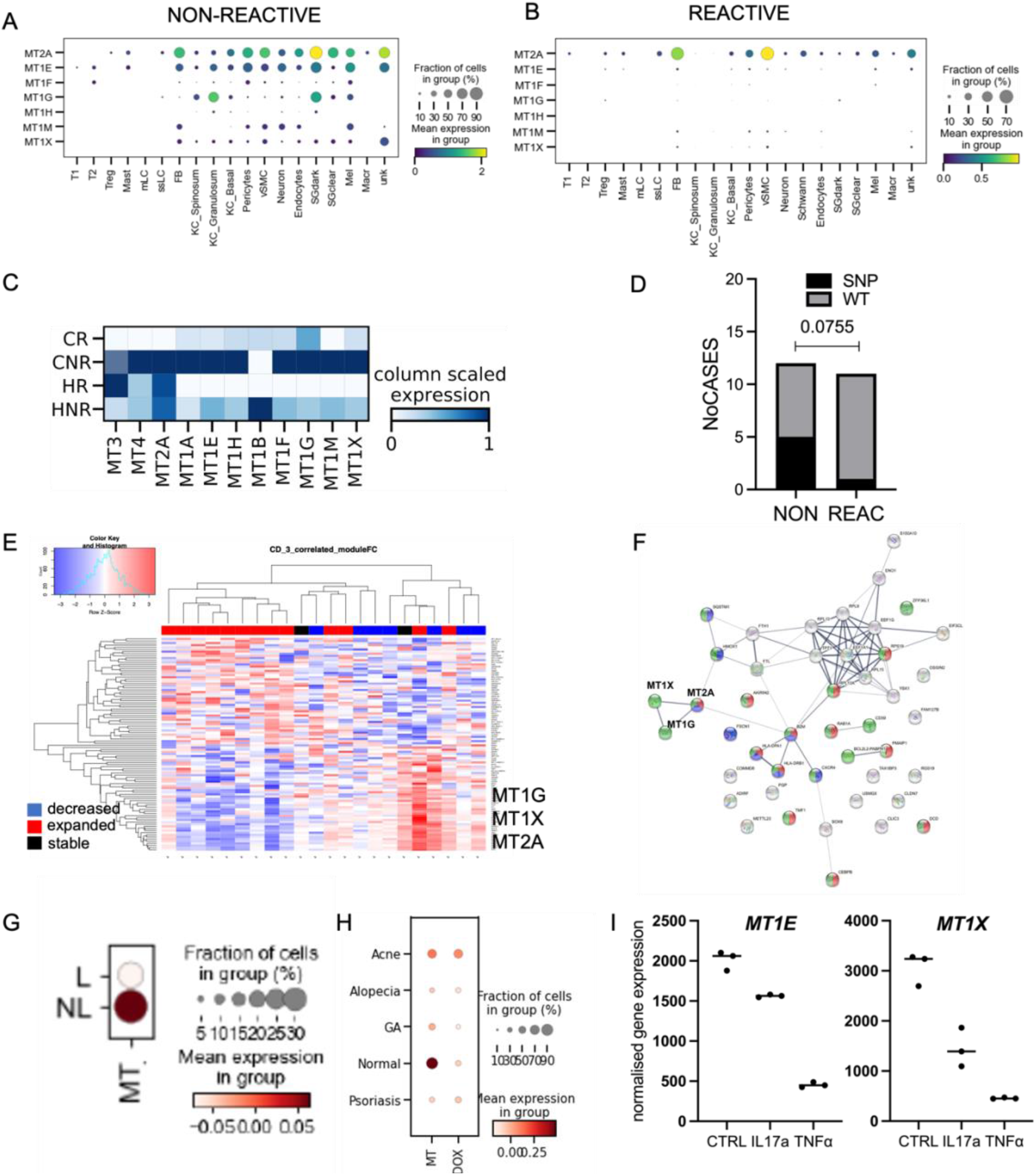
Enhanced expression of metallothionein genes protects non-reactive patients from HDM-induced T cell-mediated inflammation. A,B) Violin plots showing expression of metallothioneins across cell types in control patch tests from patients with non-reactive (A) and reactive (B) patch test reactions to HDM. SCRAN-normalised single cell RNA expression shown for each transcript. n=3, fresh skin biopsies, Drop-seq C) Heatmap comparing levels of expression for metallothioneins in whole skin using high sensitivity Constellation-seq method, n=10 skin donors. CR: control reactive, HR: HDM reactive, CNR: control non-reactive, HNR: HDM non-reactive D) Distribution of WT vs SNP in MT1X gene across patient groups E) Heatmap showing segregation of patients with CD3 T cells decreasing (blue), expanding (red) and stable (black) in reaction to patch test using fold change expression values of 100 top differentially regulated genes in module turquoise. Genes overexpressed in non-reactive samples contain members of metallothionein family. F) Protein interaction analysis (STRING) depicting metallothionein functional importance for metal transition metal ion homeostasis (red), cytokine mediated signalling (blue) and responses to stress (green) G) Expression of metallothioneins in patients with AD, in lesional (LES) and non-lesional (NLES) skin. H) Expression of signatures encoding metallothioneins (MT), and RedOX, in patients with T-cell mediated skin diseases Z-score, GSE150672 G,H) dotplot: size depict % of expressing cells, colour intensity encodes mean expression in group. Acne: Acne Vulgaris, Alopecia: Alopecia Areata, GA: Granuloma Annulare I) Normalised expression levels of genes encoding *MT1E* and *MT1X* in keratinocytes exposed to IL17a and TNFα. GSE36287.

Metallothioneins (MT) are small, cysteine-rich heavy metal-binding proteins which participate in an array of protective stress responses. Given the uniform downregulation across the skin cell types, we sought to test whether DNA polymorphisms could underpin differences in non-reactive vs reactive patients. Hypothesis driven GenePy logistic regression analysis ^33^ of 34 genes encoding oxidative stress responses, and key immunological and structural features predefined in the study (Supplementary Table 11), identified *MT1X* as the top gene differentiating patient groups. A chi-square test of independence was performed to examine the relation between single nucleotide polymorphism (SNP) in *MT1X* and reactivity to HDM. The relation between these variables was close to significant, chi^2^ = 3.96, p = 0.076, indicating that existence of a SNP in *MT1X* protected from allergen-driven inflammation (Figure 6D). Examining the genomic region of MT1X, we confirmed, that both SNPs (16:56682411:G>T and 16:56682435:C>A) were localized in the promoter and enhancer region of *MT1X* gene (Supplementary Figure 6B).

Interestingly, even though alterations in *MT1X* promoter sequence appeared to underpin reactivity to HDM, all reactive patients degraded mRNA for all type 1 metallothioneins on exposure to the allergen. Functional human metallothionein genes are clustered on human chromosome 16, and they respond to environmental stresses in a coordinated manner ^34^. This posed a possibility of existence of additional inducible regulation of metallothionein expression. WGCNA analysis pointed to a specific module of genes (turquoise, Supplementary Figure 3D, Supplementary Table 6), the expression of which varied with the level of CD3 infiltrate. Genes correlating positively with CD3 infiltrate encoded signalling receptor activity. In contrast, genes whose expression decreased with CD3+ T cell frequency encoded cellular homeostasis including response to metal ions and to cytokine stimulus (Supplementary Figure 6C, Supplementary Table 12), comprising several members of metallothionein family (*MT2A, MT1G, MT1X)*. Strikingly, LCs from samples where CD3 T cells expanded on exposure to HDM (red) were separated by reduced expression of genes in the turquoise module, including metallothioneins *MT2A, MT1G, MT1X*, ferritin chains (*FLT, FTH1*) and heme oxygenase (*HMOX1*) (Figure 6E), creating a network of antioxidant defences (Figure 6F). We next confirmed the expression of metallothionein genes was decreased in chronic AD lesions (Figure 6G) and compromised across a range of T-cell mediated skin diseases, in comparison to healthy skin (Figure 6H). Furthermore, analysis of publicly available data of keratinocytes exposed to a range of cytokines, indicate, that expression of metallothioneins can be downregulated by IL17a and TNFα (Figure 6I), providing an inducible mechanism by which T cell mediated allergic immune responses deplete the RedOx buffer, and render the skin susceptible to chronic inflammation.

## DISCUSSION

Accurate regulation of cutaneous immunity is fundamental for human health and quality of life as inappropriate immune activation results in inflammatory disorders, affecting up to 40% of the population ^35–37^. In AD this problem is particularly severe, manifested by frequent exacerbations and resulting in significant morbidity in paediatric and adult patients^6,8,9^.

Comparing cutaneous and systemic responses of eczema patients to HDM, we demonstrated that despite evident allergen-specific responses in blood, nearly 50% of patients did not react clinically to an epicutaneous patch test with allergen. We ruled out the possibility that the lack of reactivity might be due to the epidermal permeability barrier preventing penetration of the allergen, as these individuals had a less effective barrier as indicated by increased TEWL. This suggested the existence of local epidermal tolerance in the non-lesional skin and posed a question about the factors regulating the distinctly different outcomes between reactive and non-reactive patients. To explore the molecular and cellular networks underpinning immune reactiveness, we took biopsies from control and HDM-exposed sites, and subjected isolated cell populations to micro-bulk and single cell transcriptomic analysis. These were validated with protein expression measured by flow cytometry and *in situ*. We further investigated exome sequence in a subset of genes preselected based on the transcriptomic analysis, to delineate potential single nucleotide polymorphisms which might be conferring protection from allergen-driven skin inflammation. This methodological approach allowed us to contrast immunoreactivity versus tolerance as it is happening in human skin *in vivo*.

Our study documents that immunotolerance is the baseline state of cutaneous immune network in AD, similar to healthy skin ^38^, and is mediated by numerous interactions across multiple parenchymal and immune cell subsets. In allergen-responsive individuals, this innate state of tolerance is overcome by inflammatory signalling, epitomised in crosstalk between activated Tfh Th17 TNF-expressing T cells and LCs. Importantly, the baseline state in skin is distinctively different in non-reactive and reactive patients. These baseline differences are localised mainly to the epidermis and the immune compartment. In contrast, following allergen application, when the inflammatory reaction has developed and has formed a skin lesion, activation of other cell types becomes evident, similar to changes observed between non-lesional and lesion skin of AD patients ^18,20,29^. Interestingly, our analyses indicate that while sub-clinical inflammation in keratinocytes seems to pre-set the epidermal microenvironment, specific signals from mast cells and fibroblasts, including FAM3C on mast cells to CLEC2D on T cells drive TNF expression. The expansion of LC in response to HDM is intriguing; while expansion of dermal DC can be attributed to infiltration of monocytic precursors from blood, the epidermal compartment is independent of blood progenitors. Thus, the expansion would need to be due to local rapid proliferation, differentiation from skin resident precursors or local chemotaxis from epidermis surrounding the patch test site, in response to signals induced in the inflammatory patch test reaction. A similar observation was made recently when contrasting lesional and non-lesional AD skin ^29^, indicating that allergic reactivity induced by HDM mimics biological changes in chronic lesions, and suggesting LC expansion is an early event in lesion formation. Furthermore, changes we observed in expression of CD207, a key molecule for pathogen sequestration in skin, may point to a mechanism compromising anti-microbial defences in AD skin, and thus perpetuating inflammation.

The frequency and the state of activation of Tfh Th17 TNF-expressing T cells appears to be critical for subsequent reaction to the allergen. Tfh have been previously implicated in driving re-call allergic responses to HDM in lungs ^39^. However, our study defines these cells as Th17 TNF-expressing, in contrast to Th2 cells reported by others. Th17 T cells have been already shown to be of importance in AD, expressed at higher levels in skin of children than in adults ^14^, and higher in intrinsic AD ^15^. Our previous analysis of skin immunophenotypes indicated existence of a specific endotype of Th17 high patients with AD, highly prevalent in chronic lesions ^17^. The role of Th17 skin infiltrating T cells in driving acute responses to an allergen could be conceivably executed via contributing to an augmented state of immune readiness, promoting more severe immune reaction. While IL17 has been established as driving innate anti-microbial responses, TNFα is a particularly multifunctional cytokine, exerting effect on numerous skin populations. TNFα drives expression of adhesion molecules in skin of patients with AD, which may facilitate immune cell extravasation ^40^. Together with Th2 cytokines TNFα induces atopic dermatitis-like features on epidermal differentiation proteins and stratum corneum lipids in human skin equivalents ^41^. Induced by S.aureus, TNFα leads to up-regulation of HLA-DR molecules in keratinocytes and facilitates HDM allergen presentation^12^. Inflammatory IL-17 and TNFα secreting CD4(+) T cells have been shown to persist even in highly immunosuppressive cancer environments ^42^, indicating their potential to overcome mechanisms of cutaneous homeostasis.

TNFα secreted by T cells could play a double role in driving LC function by regulating both LC migration and maturation ^26,43–46^. Expressed in unperturbed skin, it can drive LC activation. On exposure to HDM, TNF produced by activated T cells will provide a chemotactic signal for LC to migrate out of the epidermis. In the hub structures observed in HDM reactive patients, activated LCs would support T cell activation and survival, perpetuating inflammation. However, in addition to delivering chemotactic and pro-maturation signals, TNFα has cytotoxic functions, likely resulting in the observed expansion and loss of functional transcriptomes of LCs. Several lines of evidence point to the importance of the ability of Langerhans cells (LCs) to induce immunotolerance as critical for cutaneous homeostasis ^38,47,48^. In this context, loss of LC function could likely lead to uncontrolled inflammation *in situ*, as observed in the HDM reactive patch test. Such exhaustion induced by overactivation has previously been observed in dendritic cells and macrophages in chronic infection and when overwhelmed by antigen load ^49,50^

In contrast to sub-clinical inflammation in the skin of reactive patients, expression of metallothioneins was associated with protection from HDM-driven inflammatory reaction. Levels of expression of metallothioneins seemed to be controlled both at the constitutive (via SNP in *MT1X* gene) and inducible levels (regulated by the acute oxidative stress/inflammation), highlighting the importance of anti-oxidative defences in AD skin. Importantly, anti-oxidative defence was one of the transcriptomic modules critically compromised in LCs from reactive patch test sites. Oxidative stress is one of the key components driving allergic sensitisation, and in asthma models, aeroallergens such as HDM, directly induce production of reactive oxygen species and DNA damage and dampen antioxidant responses ^51,52^. Additionally, we and others demonstrate that cytokine signalling can affect metallothionein expression in an inducible manner. Indeed, increased oxidative stress during AD exacerbation^53^ and decreased antioxidant capability in children with eczema^54^ has been previously observed. Since oxidative stress itself exhausts antioxidant responses and can be induced by many eczema-associated factors including allergens, hormones and chemicals, it is possible that chronic exposure to such triggers may compromise LC function in the epidermis of individuals with eczema making them less able to maintain cutaneous tolerance and extending our observation beyond the patch test system.

Based on our analyses, we propose a model in which sub-clinical inflammation exhausts metallothionein stores in the skin of genetically pre-disposed patients. This leads to the poised state of keratinocytes, expressing high levels of *MAP4K5* and other kinases. The sub-cutaneous inflammation is in parallel manifested by infiltration TNFα expressing activated T cells, and activated antigen presenting cells, including LCs. In response to allergen, these quickly initiate inflammatory responses and expand T cell population driving inflammatory reaction. This in turn leads to exhaustion of LCs, uncontrolled inflammation and lesion formation thereby mediating clinical signs of inflammation. In healthy/non inflamed skin allergen exposure, in the absence of subclinical inflammation, mediates immunological non-responsiveness, but in the inflamed skin depleted of oxidative defences, exposure to HDM initiates allergic inflammation.

Our current study provides a detailed description of cellular and molecular crosstalk in the skin of eczema patients, proposing a mechanism supporting development of allergen-induced inflammation. We conclude, that while immunotolerance conferred by LCs in healthy skin is preserved in non-lesional skin of eczema patients, this tolerance is compromised on activation of allergen-specific T lymphocytes infiltrating skin. lymphocytes infiltrating skin. Therefore, therapeutic strategies aiming to restore the healthier, tolerance-related signaling in the cutaneous immune network could help in preventing eczema exacerbations, harnessed to improving skin health and in relation to treating active skin disease lesions in patients with AD.

## MATERIALS AND METHODS

### Study design

Informed, written consent was obtained as per approval South East Coast -Brighton & Sussex Research Ethics Committee in adherence to Helsinki Guidelines (approval: 16/LO/0999). Adult AD patients with mild to severe disease (mean objective EASI) were recruited through the Dermatology Centre, Southampton. University Hospital NHS Trust. All AD patients fulfilled the diagnostic criteria for AD as defined by the UK. AD diagnostic criteria^22^.

### *In vivo* allergen challenge model

House Dust Mite allergen (ALK-Abello, Horsholm, Denmark AD01-AD10, Citeq Biologics, Netherlands AD11-AD28) has been applied in paraffin via epicutaneous patch application to the upper buttock skin at a non-lesional site (free from eczema) following 10x tape strip procedure to remove stratum corneum, according to our previous method ^1,59^. A control patch was applied in parallel following identical procedure, except for the HDM allergen. In all AD volunteers, this site showed no evidence of active eczema, and the volunteers were not being treated with topical therapy. Clinical responses were quantified 48 hours later at each challenge site by a specialist registrar in dermatology trained in patch testing. 6 mm skin biopsies were taken under local anaesthesia from allergen-exposed and control skin.

For detailed Materials and Methods: Experimental Procedures see Supplementary Materials.

## Data Availability

All data produced in the present study will be available upon reasonable request to the authors following acceptance for publication by peer review.

## Acknowledgments

We acknowledge the use of the IRIDIS High Performance Computing Facility and Flow Cytometry Core Facilities, together with support services at the University of Southampton. Authors wish to acknowledge Nikki Graham, Senior Technician within the DNA laboratory, Dr Carolann McGuire and Dr Richard Jewell from Flow Cytometry Core for technical support. We are grateful to nurses and administrative staff at the Clinical Research Facility, NIHR, University Hospital Southampton NHS Foundation Trust. We express our deepest gratitude to the patients recruited to the study.

## Funding

This research was funded in whole by the Wellcome Trust [Sir Henry Dale Fellowship 109377/Z/15/Z]. For the purpose of open access, the author has applied a CC BY public copyright licence to any Author Accepted Manuscript version arising from this submission.

## Author contributions

MEP, MAJ and HS: intellectually conceived the study,

EC, YT, SH, GR, NH, MAJ, RA, MEP: patient recruitment and clinical data acquisition

SS, AV, LD, ML, NG, RA, CL: carried out experiments

SS, AV, MEP, KC: carried out analysis and meta-analysis of bulk RNA-seq data

SS, AV, MEP, KC, JD: carried out analysis and meta-analysis of single cell RNA-seq data

ES and SE: carried out WES data analysis

SS, AV, MEP: wrote the manuscript

MAJ, PF, HS, EH, CLB: discussed, and reviewed the manuscript

## Competing interests

The Authors declare no conflict of interest

## Data and materials availability

Sequencing data for RNA-seq and scRNA-seq is stored in Gene Expression Omnibus database. The exome sequencing data underlying this article cannot be shared publicly due to ethical considerations.

## Supplementary Materials

### Supplementary Materials and Methods: Experimental Procedures

### Supplementary Tables: attached as csv files

Supplementary Table 1 Patient characteristics

Supplementary Table 2 FLG mutations

Supplementary Table 3 KC signatures

Supplementary Table 4 LC transcriptional programe at baseline

Supplementary Table 5 BioLayout modules and Gene Ontology analysis

Supplementary Table 6 WGCNA modules and Gene Ontology analysis

Supplementary Table 7 Panel of genes for Constellation-seq

Supplementary Table 8 Cluster defining marker genes

Supplementary Table 9 DEGs in KC Constellation-seq

Supplementary Table 10 DEGs in all cells : contrast in HDM exposed samples

Supplementary Table 11 Gene signatures for GWAS

Supplementary Table 12 WGCNA module Turquoise: Top 100 genes

### Supplementary Materials and Methods: Experimental Procedures

#### Study Design

Objective EASI was measured as described previously^55^. Before sampling, patients were washed out from any immunosuppressive treatment for at least 5 half-lives of the drug. Atopy status was assessed for each patient using Skin Prick Test (SPT) to six most common allergens: house dust mite, grass pollen, tree pollen mix, mixed mould, cat and dog plus histamine as a positive control (ALK-Abello, Horsholm, Denmark). Maintenance of normal epidermal barrier function was measured by trans-epidermal water loss (TEWL). On enrolment, information about participants’ demographics and previous medical history, immediate family history and information about atopic disease (eg eczema, rhinitis) in the subject was collected based on the ISAAC questionnaire ^56^. Peripheral blood mononuclear cells (PBMC) were separated from venous blood and processed for DNA extraction. FLG mutation analysis was performed as described previously ^57,58^. Briefly, primer pairs were used to amplify the region of interest from DNA prepared from peripheral blood samples of individuals with atopic dermatitis and controls. FLG variants R501X, 2282del4, S3247X and R2447X, which covers more than 90% of FLG mutations in a UK population, were then identified using restriction enzyme digest of PCR products with agarose gel electrophoresis and FLG variants were confirmed by whole exome sequencing.

#### Cell isolation

6 mm biopsies were minced using a surgical scalpel and digested for 16h at 37C with agitation in RPMI with LiberaseTM (Roche) following manufacturer’s instructions. After 16h of digestion cells were collected and washed with RPMI 5% FBS. Cells were resuspended in PBS 1% BSA 20mM EDTA and filtered through 70um sterile filters before surface antibody staining for FACS or processing for Dropseq analysis.

#### Flow cytometry and cell sorting

All antibodies were used at pre-titrated, optimal concentrations. For surface staining of live cells buffer containing PBS 1% BSA was used for all antibody staining. FACS Aria flow cytometer (Becton Dickinson, USA) was used for analysis of human LCs for the expression of CD207, CD1a, HLA-DR (mouse monoclonal antibodies, CD1a, CD207:Miltenyi Biotech, UK and HLA-DR: BD Biosciences, UK) or T cells for the expression of CD3, CD25 and CD103 (Miltenyi Biotech). Freshly isolated PBMCs were activated with anti-CD3 and anti-CD28 (1 mg/μ1), with GolgiPlug (BD Biosciences, Oxford, UK). The Cytofix/Cytoperm kit (BD Biosciences) was used according to the manufacturers’ instructions.

Flow cytometric analysis with the FACS Aria flow cytometer (BD Biosciences) was undertaken following lymphocyte gating on Forward/Side scatter. Subsequent gating on CD3-PerCP 5.5 (eBiosciences), CD4-VioGreen (Miltenyi Biotech) was based on appropriate negative controls to demonstrate IL13-APC and IL-17-FITC positive cells (Miltenyi Biotech) for analysis with the FlowJo software (Tree Star, Ashland).

Singlets (FCSA:FCSH), CD207+/CD1a+ digested LCs were sorted into trizol for RNA isolation. In parallel, LC-depleted skin cells were sorted into RPMI 5% FBS and processed for immediate cryostorage.

#### Immunofluorescence microscopy of frozen tissue sections

Snap frozen skin samples were embedded in OCT (CellPath) and cut to 5-10 µm cryosections onto APES-coated slides. Sections were fixed in 4% paraformaldehyde, washed with PBS, blocked with PBS + 1% BSA + 10% FBS and incubated for 30 minutes with primary antibodies to the following markers: Langerin (Leica), multi-cytokeratin (Leica), CD3 (Dako), CD4 (Abcam), CD8 (Abcam) or FOXP3 (Abcam). After washing off the primary antibodies, secondary antibodies were added; these included: Alexa Fluor 488 goat anti-mouse IgG1a, Alexa Fluor 555 goat anti-rabbit IgG, and Alexa Fluor 647 goat anti-mouse IgG2b (all from ThermoFisher Scientific). Sections were then counterstained with DAPI (Sigma), mounted with Mowiol (Harco), coverslipped and imaged using an Olympus Dotslide scanning fluorescence microscope and Olympus VS-Desktop software.

#### RNA-seq

RNA was isolated using Direct-zol RNA micro prep (Zymo, UK) as per the manufacturer’s protocol. RNA concentration and integrity was determined with an Agilent Bioanalyser (Agilent Technologies, Santa Clara, CA. Preparation of RNA-seq libraries and sequencing were carried out by Source Bioscience, UK. cDNA libraries were generated using SMART-Seq Stranded Library Preparation for Ultra Low Input according to the SMART-Seq Stranded Kit User Manual following the Ultra low input workflow (Takara Bio). Samples were pooled (12/batch) for library preparation. Amplified libraries were validated on the Agilent BioAnalyzer 2100 to check the size distribution and on the Qubit High Sensitivity to check the concentration of the libraries. All the libraries passed the QC step. Sequencing was done on Illumina HiSeq 4000 instrument, 75bp PE runs, 20 ×10^6^ reads per sample.

#### Drop-seq

Freshly dissociated whole skin biopsies were suspended in RNAse-out buffer and processed on ice to the co-encapsulation of single cells with genetically-encoded beads (Drop-seq, ^60^). Monodisperse droplets at 1 nl in size were generated using the microfluidic devices fabricated in the Centre for Hybrid Biodevices, University of Southampton. To achieve single cell/single bead encapsulation with barcoded Bead SeqB (Chemgenes, USA), microfluidics parameters (pump flow speeds for cells and bead inlets, cell buoyancy) were adjusted to optimise cell-bead encapsulation and the generation of high quality cDNA libraries. Based on encapsulation frequencies and bead counts up to 2000 STAMPS /sample were taken further for library prep (High Sensitivity DNA Assay, Agilent Bioanalyser, 12 peaks with the average fragment size 500 bp). The resulting libraries were run on a shared NextSeq run (4×10^4^ reads/cell for maximal coverage) at the Wessex Investigational Sciences Hub laboratory, University of Southampton, to obtain single cell sequencing data

#### Constellation-seq

Single cell libraries were generated using the Chromium Single Cell 3’ library and gel bead kit v3.1 from 10x Genomics. Briefly, cell suspensions were tagged using TotalSeq™ hashtag antibodies (Biolegend). After pooling, 10,000 viable cells were loaded onto a channel of the 10x chip to produce Gel Bead-in-Emulsions (GEMs). This underwent reverse transcription to barcode RNA before clean-up and cDNA amplification. cDNA was used for targeted linear amplification comprising 20 rounds of linear amplification (60°C) using a pool of primers (Supplementary Table 7) at 40 nM and 0.4 µM of a P5 3’blocked primer as described previously(Vallejo et al). cDNA libraries were purified twice using AMPure XP (Beckman Coulter) magnetic beads (1:0.6) and libraries assessed using a Bioanalyser before tagmentation and Next-seq sequencing on an Illumina Nextseq500, (paired end 28×60 bp reads).

#### Bulk RNA-seq data analysis

Quality control for FASTQ files with raw sequence data was done using FASTQC tool [FastQC: a quality control tool for high throughput sequence data. Available online at: http://www.bioinformatics.babraham.ac.uk/projects/fastqc]. High-quality reads filtered at 15M depth across all samples were mapped to the human genome (GRCh38) using Kallisto^61^. Raw counts from RNA-Seq were processed in Bioconductor package EdgeR^62^ and SLEUTH^63^, variance was estimated and size factor normalized using trimmed mean of M-values (TMM). Genes with minimum 2 reads at minimum 50% samples were included in the downstream analyses. Differentially expressed genes (DEG) we identified applying significance threshold FDR p<0.05, |LogFC|>1. Normalised reads were taken for transcript-to-transcript co-expression analysis (BioLayout ^64^). Pearson correlation coefficient r=0.85, Markov Clustering Algorithm = 1.7.. WGCNA analysis ^27^were run on 5000 genes with maximum median absolute deviation (MAD), at power=4 module size 30 in R v 4.0.3 using voom transformed (TMM) normalised expression data. Gene ontology analysis across clusters and modules was done using ToppGene online tool^65^.

#### scRNA-seq data and Constellation-seq analysis

Single cell RNA-seq and Constellation-seq analysis was carried out using pipelines established in Systems Immunology Group ^26,28^. Following demultiplexing, raw FASTQ files were aligned to the human genome (GRCh38), using kallisto-bustools ^61^. CellBender^66^ was used to remove empty technical artefacts. Doublet detection and hashing demultiplexing was done using Solo ^67^. Visualization and clustering of scRNAseq was performed in Scanpy ^31^ following standard quality checks (empty barcodes, percentage of mitochondrial genes). Clusters were determined via single-cell neighbourhood analyses on first principal components followed by clustering and cell type identification (leiden-based clustering ^68^. Cell types were annotated based on the expression of known marker genes, and cross-validated using publically available single cell transcriptomes ^29^. Differentially regulated transcriptional networks were identified using model-based analysis of single-cell transcriptomics MAST ^69^. Specific transcriptional signatures were tracked using Gene Set Expression Analysis ^70,71^. Transcription factor activity prediction was done using DoRothEA^72^. Cell-cell communication was inferred using CelphoneDB^73^ and CosstalkR^74^.

#### Whole Exome Data generation

Whole exome sequencing was performed by Macrogen, with data uploaded to the University of Southampton supercomputer IRIDIS5 in February 2021. Agilent SureSelect Human All Exon V6 capture kit was used for all 28 samples.

#### Whole exome data analysis

The raw fastq files were aligned to the human genome reference GRCh38 with additional HLA regions included. Alignment was performed using BWA-MEM v0.7.15-r1140 and Samtools ^75^ v1.3.1. Picard (http://broadinstitute.github.io/picard/) was used to mark duplicates, sort the BAM files, index, and fix the mate pairs. GATK^76^ v4 base quality score recalibration (BQSR) was used to detect systematic errors by the sequencing machine when it estimates the accuracy of each base call. Joint-calling was executed using a bespoke script. GATK GenomicsDBImport was used to create a database of all g.vcf files for joint-calling and was targeted to the intersection between Agilent SureSelect V6 and Agilent SureSelect V5, both with +/-150bp padding. GATK Genotype GVCF was applied to joint-call the 28 AD samples with 1100 inflammatory bowel disease patients. A final script applied GATK Variant Quality Score Recalibration (VQSR), a technique applied on the variant callset that uses machine learning to model the technical profile of variants in a training set and uses that to flag probable artefacts from the callset. Annotation was completed using Ensembl VEP^77^ v103. The joint-called vcf was uploaded to a local installation of seqr (https://github.com/broadinstitute/seqr) on a virtual machine for data visualisation, analysis, filtering and reporting. A suite of bespoke scripts was used to assess quality control. Bedtools^78^ v2.26.0 was applied to calculate exome data coverage relative to the target capture kit. GATK VariantEval tool was used to calculate various quality control metrics including: the number of raw or filtered SNPs and the ratio of transitions to transversions. These metrics are further stratified by functional class, CpG site, and amino acid degeneracy. Picard CollectVariantCallingMetrics was applied to collect the per-sample and aggregate (spanning all samples) metrics from the provided vcf file. Peddy (https://github.com/brentp/peddy) was executed locally in a python conda environment to assess the relatedness of individuals, and predict their ancestry and sex. All QC metrics data were compiled into a Shiny App using R v4.2.0 for data visualisation.

#### Identifying filaggrin variants

We selected all loss of function filaggrin (FLG) variants with a maximum allele frequency (across all populations in gnomAD) of <0.05, allele balance of >0.2 and genotype quality > 0.3 with variants in FLG, with VQSR applied as a flag.

#### Applying GenePy to identify genes enriched in reactive vs non-reactive patients

We assessed the difference in gene mutation burden between patients non-reactive and reactive to HDM using GenePy 1.3 (Mossotto et al). We initially calculated GenePy scores for a pre-selected 34 genes (Supplementary Table 11) and compared their scores between non-reactive and reactive patients using logistic regression. We extrapolated this analysis to 2002 autoimmune genes (Supplementary Table 11) and compared GenePy scores using a Mann-Whitney-U test.

#### Statistical analysis

All statistical analysis were carried out using GraphPad Prism V9.2.0 unless specifically state otherwise.

#### Supplementary Figures

**Supplementary Figure 1.**
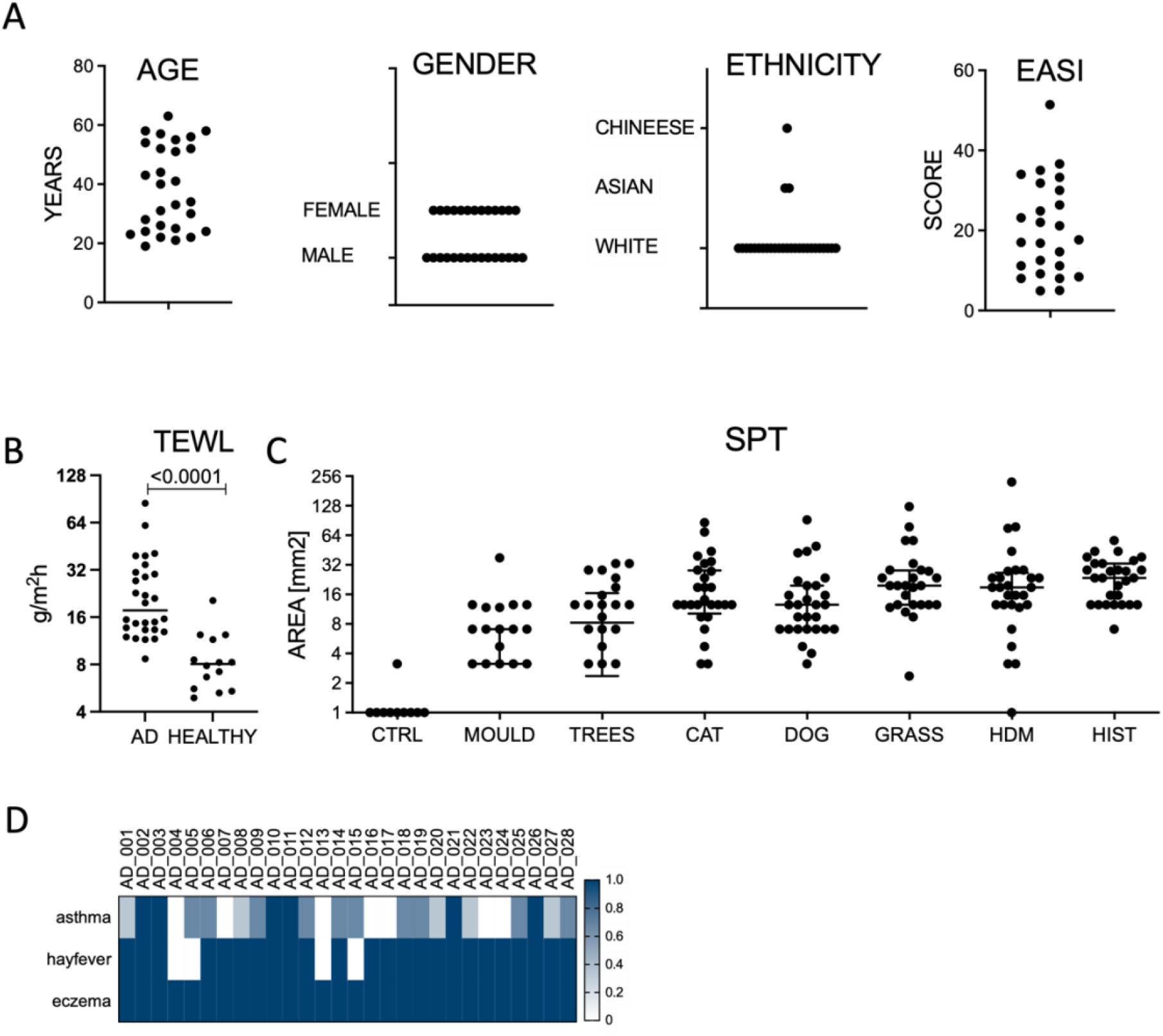
*In vivo* allergen challenge model to investigate mechanisms of local immune responses in human skin. Patient cohort characteristics: A) age, gender, ethnicity, EASI score, n=28. B) B) trans-epidermal water loss (TEWL) in patients with AD and healthy controls, C) Skin Prick Test (SPT) responses to stimulation with 6 most common allergens (wheal area given). C) Recorded responses to ISAAC questionnaire, 1 represents 100% of positive responses to questions in a category.

**Supplementary Figure 2.**
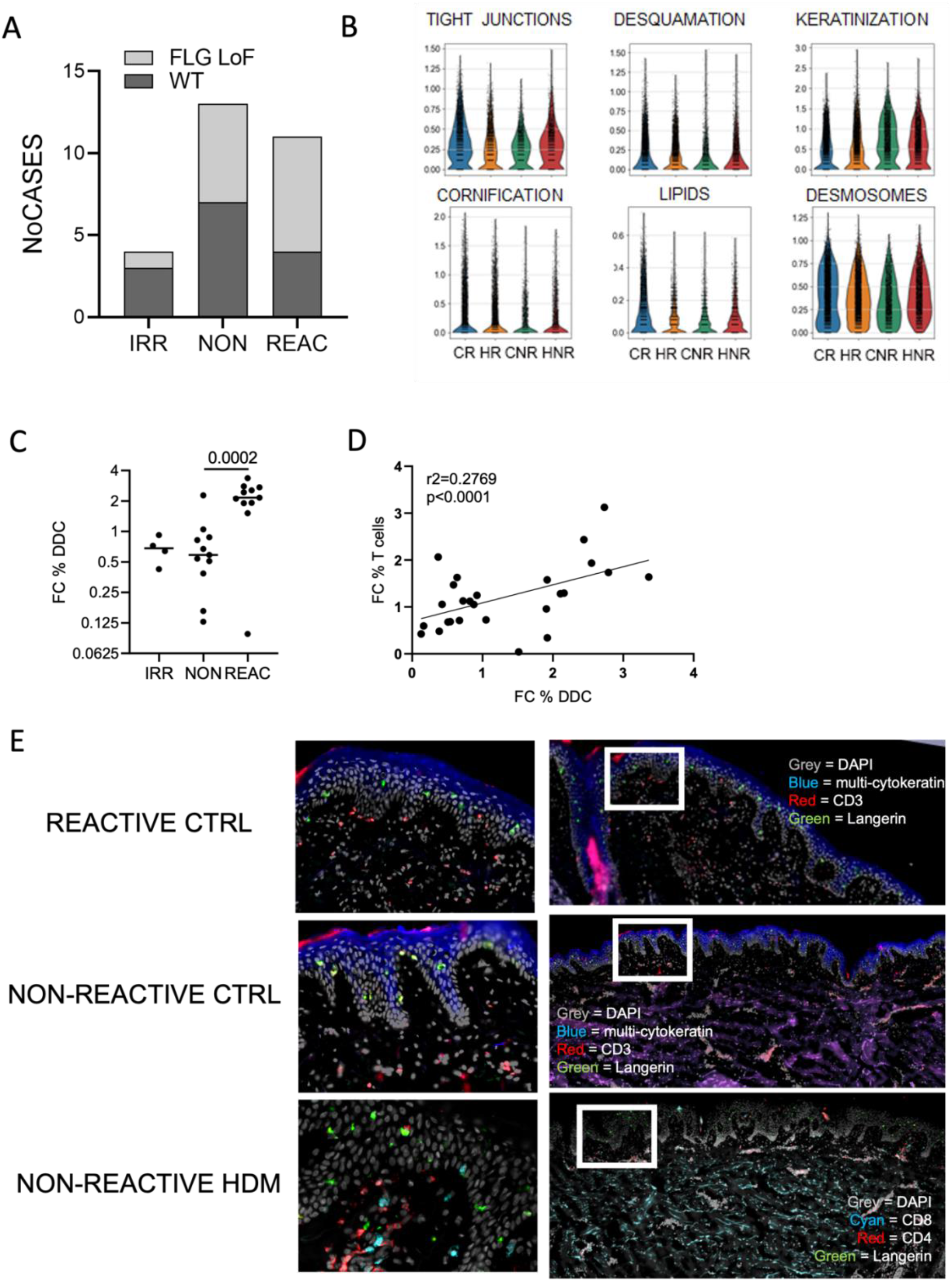
Reactivity to HDM is mediated by co-expansion of T cells and LCs. A) Number of irritant (IRR), non-reactive (NON), and reactive (REAC) cases with loss of function (LoF) variants in FLG compared to wildtype (WT). 7 SNP measured using Whole Genome Sequencing, n=27 patients. B) Z scores for keratinocyte gene expression programmes, DropSeq whole transcriptome analysis, fresh tissue, n=6 paired biopsies from 3 donors CR: control reactive, HR: HDM reactive, CNR: control non-reactive, HNR: HDM non-reactive C) Fold changes in percentage of detected DDC between HDM patch test and control patch test from patients with irritant, non-reactive and reactive reactions to HDM. Statistical significance assessed by t-test D) Correlations between fold changes in percentage of CD3+ T cells and DDCs. Pearson correlation coefficient shown. N) Immunofluorescence staining of patch test sites from reactive and non-reactive patients. Inserts show the indicated optical fields. CD207 (green) and CD3 (red). Epidermal layer stained with multi-cytokeratin (blue). DAPI stain for nuclei (grey). In patch test from HDM exposed non-reactive patient example of interaction between CD4 (red) and CD8 (cyan) T cells shown.

**Supplementary Figure 3.**
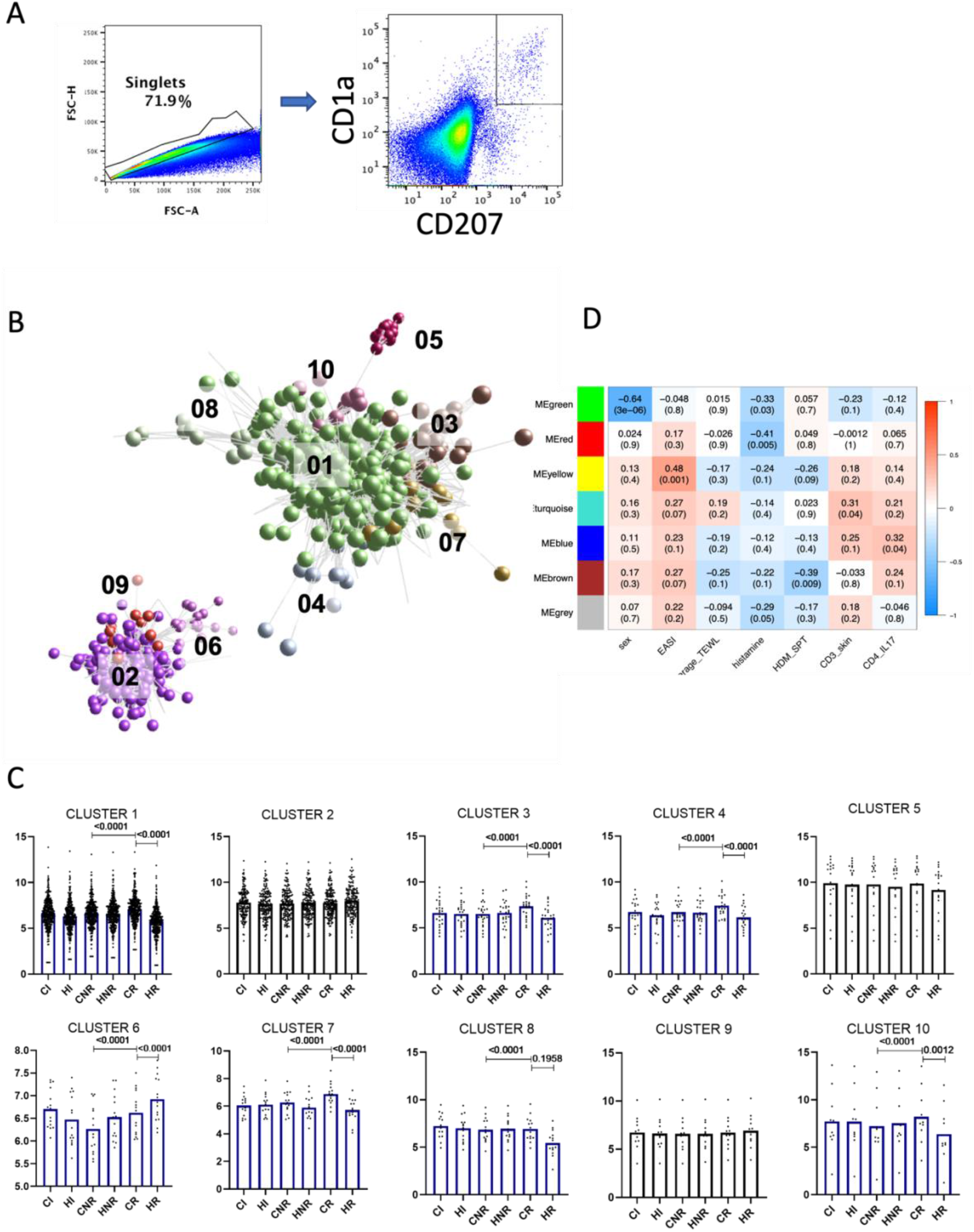
HDM reactivity in reactive patients is mediated by LC : T cell TNF crosstalk and impairs LC transcriptional programming. A) Gating strategy for sorting CD207+CD1a+ Langerhans cells, B) Transcript-to-transcript correlation network of 10 largest clusters detected in LC transcriptomes. Lines (edges) represent the similarity between transcripts, circles (nodes) represent genes. C) Mean (± SEM) expression profiles for clusters 1-10, across sample groups. CI: control irritant, HI: HDM irritant, CR: control reactive, HR: HDM reactive, CNR: control non-reactive, HNR: HDM non-reactive. Dots represent average expression of transcript. Statistical comparison done using Mann Whitney test for paired samples. D) WGCNA analysis of correlation between clinical and experimental features, recorded as continuous variables and eigenvectors representing modules of co-expressed genes across LC transcriptomes. Each colour coded module (left) represents co-expressed genes. Clinical/experimental features are labelled across X axis. Person correlation coefficients and p-values are given for each correlation, the heatmap (blue to red) indicates correlation strength and direction.

**Supplementary Figure 4.**
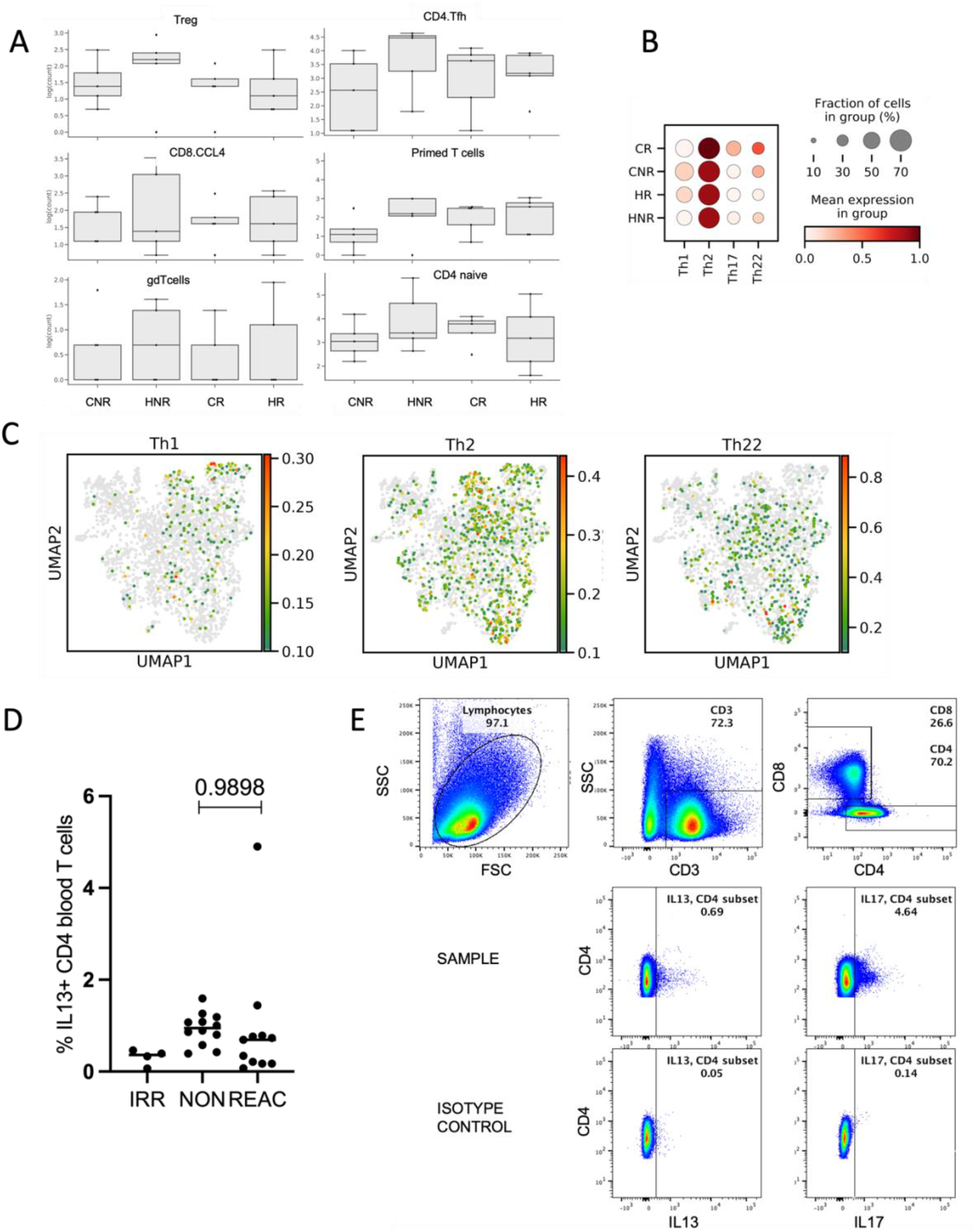
Activated TNF-expressing Th17 cells are significantly enriched at baseline in reactive patients. A) Composition of T cell subsets (HCA profiles) across patch test phenotypes. CR: control reactive, HR: HDM reactive, CNR: control non-reactive, HNR: HDM non-reactive. B) Th immunotypes gene signature across patient groups, dotplot: size depict % of expressing cells, colour intensity encodes mean expression in group C) %IL13 producing CD3+CD4+ Tcells from PBMCS in non-reactive (NRC) and reactive (R) patients. T-test. C) UMAP plot of 2374 single T lymphocytes cells. Constellation-seq analysis enriched for 1161 transcripts from patch test skin biopsies, n=10 patients, 5 per group. Th immunotypes for T cell polarisation shown (Z-score, green to red). D) Gating strategy for flow cytometry analysis of intracellular cytokine staining.

**Supplementary Figure 5.**
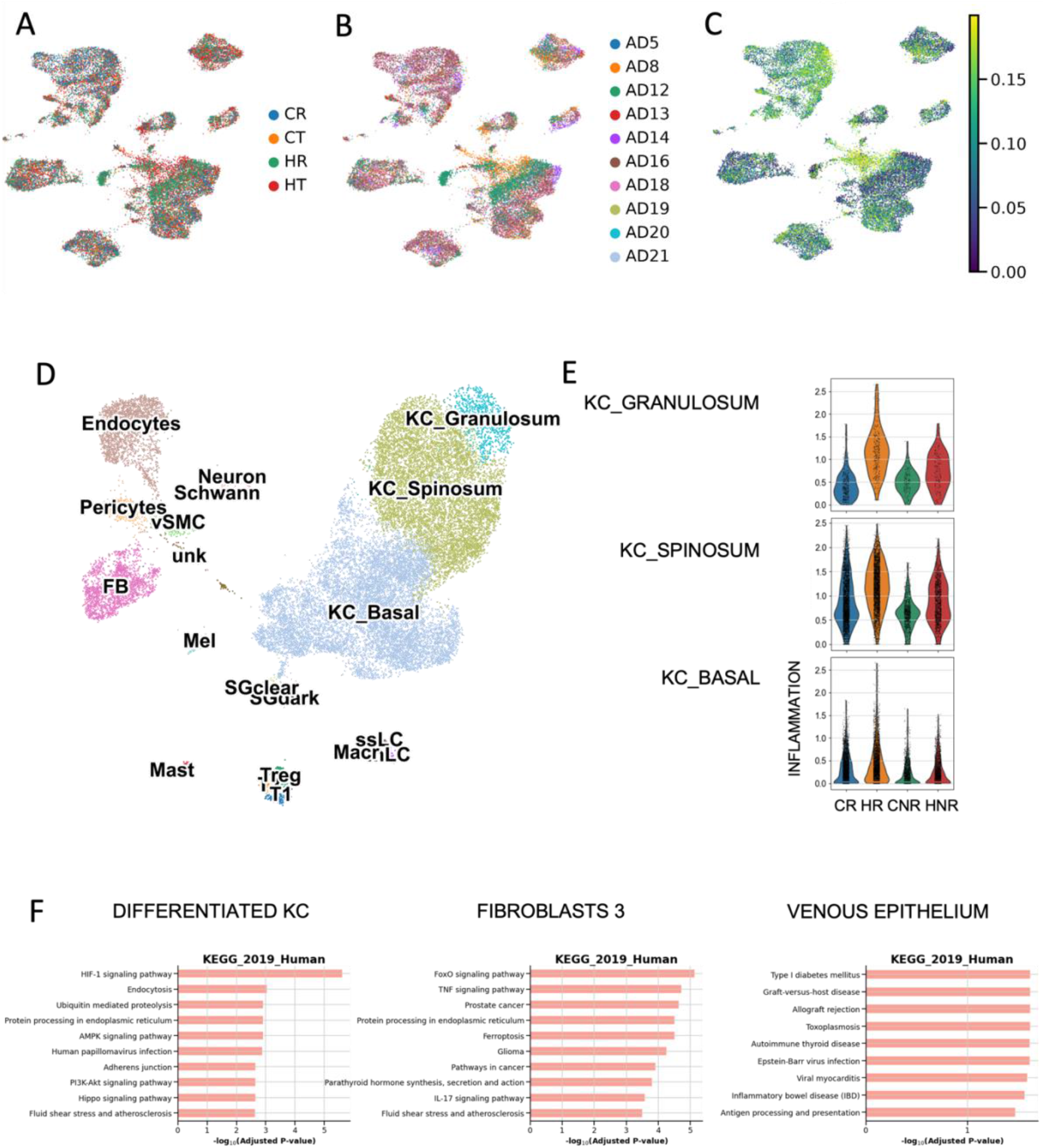
Sub-clinical inflammation and environmental stress in differentiated keratinocytes poise immune status of reactive patients. A-C) Quality controls for constellation-seq analysis enriched for 1161 transcripts in 25844 single cells from patch test skin biopsies, n=10 patients A) BBKNN integration across treatment and condition B) BBKNN integration across donors C) percentage of mitochondrial genes across samples UMAP plot depicting clustering of specific cell populations B) Cell subset defining markers (Wilcoxon rank test) D-E) DropSeq analysis of whole transcriptome across single cells isolated from fresh whole skin biopsies. N=6 paired samples, 3 donors. D) UMAP plot depicting clustering of specific cell populations E) Violin plots showing Z score of KC inflammation signature across skin layers, CR: control reactive, HR: HDM reactive, CNR: control non-reactive, HNR: HDM non-reactive F) Gene Ontology enrichment in DEGs identified across cell populations in Constellation seq.

**Supplementary Figure 6.**
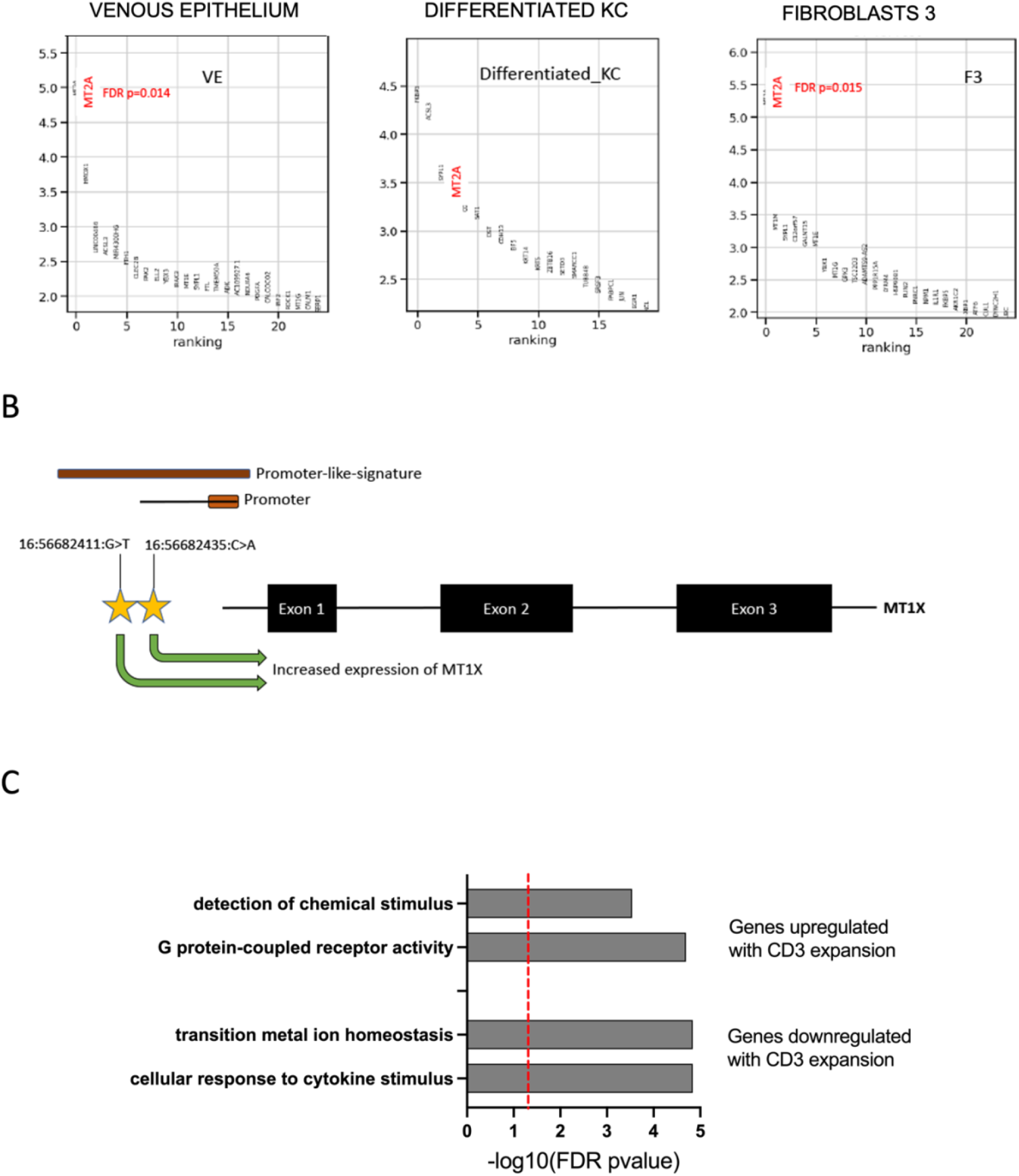
Enhanced expression of metallothionein genes protects non-reactive patients from HDM-induced T cell-mediated inflammation. A) Constellation-seq analysis, DEGs overexpressed in non-reactive vs reactive patients B) Schematic depicting localisation of SNP in promoter/enhancer region upstream of MT1X, significantly enriched in patients tolerant to HDM C) Processes significantly enriched in module turquoise, correlating with CD3 infiltrate in skin. Top processed up-regulated (blue) and downregulated (orange) with CD3 infiltration shown. Red line denotes significance cut-off by FDR p value.

## Notes

### Competing Interest Statement

The authors have declared no competing interest.

### Author Declarations

South East Coast - Brighton & Sussex Research Ethics Committee in adherence to Helsinki Guidelines gave ethical approval for this work (approval: 16/LO/0999)

